# Use of an EHR to inform an administrative data algorithm to categorize inpatient COVID-19 severity

**DOI:** 10.1101/2021.10.04.21264513

**Authors:** Elizabeth M. Garry, Andrew R. Weckstein, Kenneth Quinto, Tamar Lasky, Aloka Chakravarty, Sandy Leonard, Sarah E. Vititoe, Imaani J. Easthausen, Jeremy A. Rassen, Nicolle M. Gatto

**Affiliations:** Aetion, Inc., New York, NY, USA; Office of Medical Policy, Center for Drug Evaluation and Research, U.S. Food and Drug Administration, Silver Spring, MD, USA; Office of the Commissioner, U.S. Food and Drug Administration, Silver Spring, MD, USA; Partnerships and RWD, HealthVerity, Philadelphia, PA, USA

**Keywords:** COVID-19, severity of illness index, administrative claims, healthcare, electronic health records

## Abstract

**Importance:** Algorithms for classification of inpatient COVID-19 severity are necessary for confounding control in studies using real-world data (RWD).

**Objective:** To explore use of electronic health record (EHR) data to inform an administrative data algorithm for classification of supplemental oxygen or noninvasive ventilation (O2/NIV) and invasive mechanical ventilation (IMV) to assess disease severity in hospitalized COVID-19 patients.

**Design:** In this retrospective cohort study, we developed an initial procedure-based algorithm to identify O2/NIV, IMV, and NEITHER O2/NIV nor IMV in two inpatient RWD sources. We then expanded the algorithm to explore the impact of adding diagnoses indicative of clinical need for O2/NIV (hypoxia, hypoxemia) or IMV (acute respiratory distress syndrome) and O2-related patient vitals available in the EHR. Observed changes in severity categorization were used to augment the administrative algorithm.

**Setting:** Optum de-identified COVID-19 EHR data and HealthVerity claims and chargemaster data (March – August 2020).

**Participants:** Among patients hospitalized with COVID-19 in each RWD source, our motivating example selected dexamethasone (DEX+) initiators and a random selection of patients who were non-initiators of a corticosteroid of interest (CSI-) matched on date of DEX initiation, age, sex, baseline comorbidity score, days since admission, and COVID-19 severity level (NEITHER, O2/NIV, IMV) on treatment index.

**Main Outcome and Measures:** Inpatient COVID-19 severity was defined using the algorithms developed to classify respiratory support requirements among hospitalized COVID-19 patients (NEITHER, O2/NIV, IMV). Measures were reported as the treatment-specific distributions of patients in each severity level, and as observed changes in severity categorization between the initial procedure-based and expanded algorithms.

**Results:** In the administrative data cohort with 5,524 DEX+ and CSI- patient pairs matched using the initial procedure-based algorithm, 30% were categorized as O2/NIV, 5% as IMV, and 65% as NEITHER. Among patients assigned NEITHER via the initial algorithm, use of an expanded algorithm informed by the EHR-based algorithm shifted 54% DEX+ and 28% CSI- to O2/NIV, and 2% DEX+ and 1% CSI- to IMV. Among patients initially assigned O2/NIV, 7% DEX+ and 3% CSI- shifted to IMV.

**Conclusions and Relevance:** Application of learnings from an EHR-based exploration to our administrative algorithm minimized treatment-differential misclassification of COVID-19 severity.

## 1. INTRODUCTION

Since coronavirus disease 2019 (COVID-19) emerged in the United States (US), there have been over *42.9 million cases* ***{WHO 2020}*** with COVID-related hospitalizations at an estimated cumulative incidence over *676.4 per 100,000* persons and COVID-19 related deaths exceeding *688,000* as of October 1, 2021 ***{CDC 2020}***. Patients hospitalized for COVID-19 may have respiratory manifestations ranging from mild dyspnea to acute respiratory failure, septic shock, and/or multiple organ dysfunction ***{NIH 2021}***. Respiratory support including supplemental oxygen (O2) delivery with or without noninvasive ventilation (NIV) and invasive mechanical ventilation (IMV) may be life-saving for patients experiencing moderate-to-severe disease. Guidelines recommend maintaining blood oxygen saturation (SpO2) of 92-96% ***{NIH 2021; Barrot 2020; Chu 2018}***. High-flow oxygen via nasal cannula or loose-fitting mask at a titrated flow rate is preferred over noninvasive positive pressure ventilation and conventional oxygen therapy ***{NIH 2021; Ni 2018}***. IMV, which is indicated for patients receiving oxygen support with a low ratio of arterial oxygen partial pressure to fractional inspired oxygen (*P*aO2/*F*IO2 ≤ 200), may be initiated among patients who continue respiratory deterioration ***{Rochwerg 2017; Fan 2018}***. According to NIH guidelines ***{NIH 2021}***, ventilator maintenance among COVID-19 patients should follow standard guidelines for management of hypoxemic respiratory failure due to other causes ***{Papazian 2019}***. In more severe cases when organs start to fail, additional support may be added to IMV to help the heart and lungs pump oxygen into the blood (extracorporeal membrane oxygenation, ECMO), help the kidneys with filtration (renal replacement therapy), or improve blood and oxygen delivery to vital organs (vasopressors). Despite the increasing availability of vaccines, determining the effectiveness of potential COVID-19 therapeutics continues to be an urgent concern. As indicators of COVID-19 severity, baseline O2, NIV, and IMV respiratory support requirements are critical measures of risk, prognosis and outcomes. This was made clear in the United Kingdom RECOVERY trial, which reported lower 28-day mortality for dexamethasone (DEX) versus usual care among patients with O2 or NIV (risk ratio: 0.82, 95% confidence interval: 0.72-0.94) and IMV (0.64, 0.51-0.81), but no discernable difference among patients receiving neither (1.19, 0.91-1.55) ***{RECOVERY 2020}***. The US Food and Drug Administration (FDA) has since issued guidance recommending that patients be classified according to baseline disease severity in all studies determining the effectiveness of new COVID-19 treatments and prevention ***{FDA 2021}***. Therefore, we sought to define COVID-19 severity for an inpatient comparative effectiveness study of DEX+ versus matched non-users of corticosteroid of interest (CSI-; dexamethasone, methylprednisolone, prednisone, hydrocortisone) at the time of match (i.e., the DEX+ treatment initiation day) using an administrative real-world data (RWD) source (protocol posted to clinicaltrials.gov; NCT04926571). Matching treated and untreated patients on the day of inpatient treatment initiation requires the ability to classify severity at study entry.

The World Health Organization (WHO) developed a Clinical Progression Scale to classify COVID-19 severity ***{WHO Working Group 2020}***. While this scale was developed for determining patient outcomes, it can also be used to determine baseline severity. However, it relies heavily on the availability of clinical information that may not always be available in RWD. The FDA Sentinel Initiative also developed a practical severity score to classify patient severity ranging from asymptomatic to critical using RWD ***{Yih 2020}***. However, the categories rely on day-level diagnoses that are often unavailable or under-recorded within inpatient data sources. There is a clear need for additional mechanisms, including algorithms to determine COVID-19 severity using RWD, especially in the inpatient setting. We therefore developed a modified version of the WHO Scale that was influenced by the FDA Sentinel score to determine O2/NIV and IMV respiratory support requirements. We started with a procedure-based initial algorithm. Given its invasiveness and cost, we assumed IMV would be fairly well recorded in administrative (billing based) RWD. We had concerns, however, that O2 and NIV procedures may not be as well captured.

This paper is a result of a research collaboration agreement with the FDA to use RWD to advance the understanding and the natural history of COVID-19 in specific patient populations, as well as treatment and diagnostic patterns during the COVID-19 pandemic. The study aimed to use an electronic health record (EHR) RWD source to develop an expanded algorithm to improve upon our initial algorithm applied to administrative data for categorization of O2/NIV and IMV in patients hospitalized for COVID-19. While the motivating example was the comparative effectiveness of DEX+ versus CSI-in HealthVerity data, we aimed to develop an algorithm for an inpatient score with broad utility beyond this example to other EHR, administrative claims, and hospital billing data sources.

## 2. MATERIALS AND METHODS

### 2.1. Data Sources

Our study used two US RWD sources (see Appendix C for additional detail). First, HealthVerity data comprises medical and pharmacy open claims (sourced in near-real-time from practice management systems, billing systems and claims clearinghouses) and closed claims (sourced from insurance providers and payers), laboratory test history and results, and chargemaster administrative hospital billing data for inpatient and outpatient encounters from all US states. The data include all major payer types (commercial, Medicaid and Medicare). Second, Optum de-identified COVID-19 EHR data includes patients with suspected COVID-19, sourced from medical and administrative encounters from hospitals, emergency departments, outpatient centers, and laboratories from across the US that include diagnosis data, laboratory data with results, procedures, vital sign measurements, prescriptions written, and medications administered. There has been scientific publication of COVID-19 research using both the HealthVerity ***{Burn 2020; Gordon 2020; Harvey 2021; Murk 2020}*** and Optum ***{Hughes 2020}*** datasets.

In the medical claims of both datasets, procedures are captured via current procedural terminology (CPT), healthcare common procedure coding system (HCPCS), and international classification of disease 10th revision procedural classification system (ICD-10-PCS) codes, diagnoses are captured via ICD-10, clinical modification (ICD-10-CM) codes, and O2 or related supplies indicating an O2 procedure may additionally be captured via revenue codes. In the chargemaster data of HealthVerity, vendor charge codes also capture procedures and diagnoses; although procedures are captured daily while hospitalized, diagnoses are primarily captured at discharge, with some further capture of diagnoses present at admission. In contrast, the Optum de-identified COVID-19 EHR has day-level diagnosis encounters while hospitalized. Comparing the available data in these two data sources offers the opportunity to create a more widely-applicable COVID-19 severity score that can be used in EHR, claims and/or hospital billing data.

### 2.2. COVID-19 severity levels

The WHO Clinical Progression Scale scores range of COVID-19 severity outcomes from uninfected (score of 0) to dead (10; see Table 1) ***{WHO Working Group 2020}***. We developed a modified version, referred to here as the mWHO score, that restricts to severity levels applicable to hospitalized COVID-19 patients (WHO original scores of 4-9), collapsed into three mutually exclusive categories for neither O2/NIV nor IMV, O2 or NIV without IMV, and any IMV with or without additional support (NEITHER, O2/NIV, and IMV, respectively; *see Appendix A for additional mWHO algorithm detail*). These categories correspond to the FDA Sentinel’s Moderate, Severe, and Critical categories, respectively, leaving out the Asymptomatic and Mild categories that apply only to non-hospitalized patients ***{Yih 2020}***.

**Table 1.**
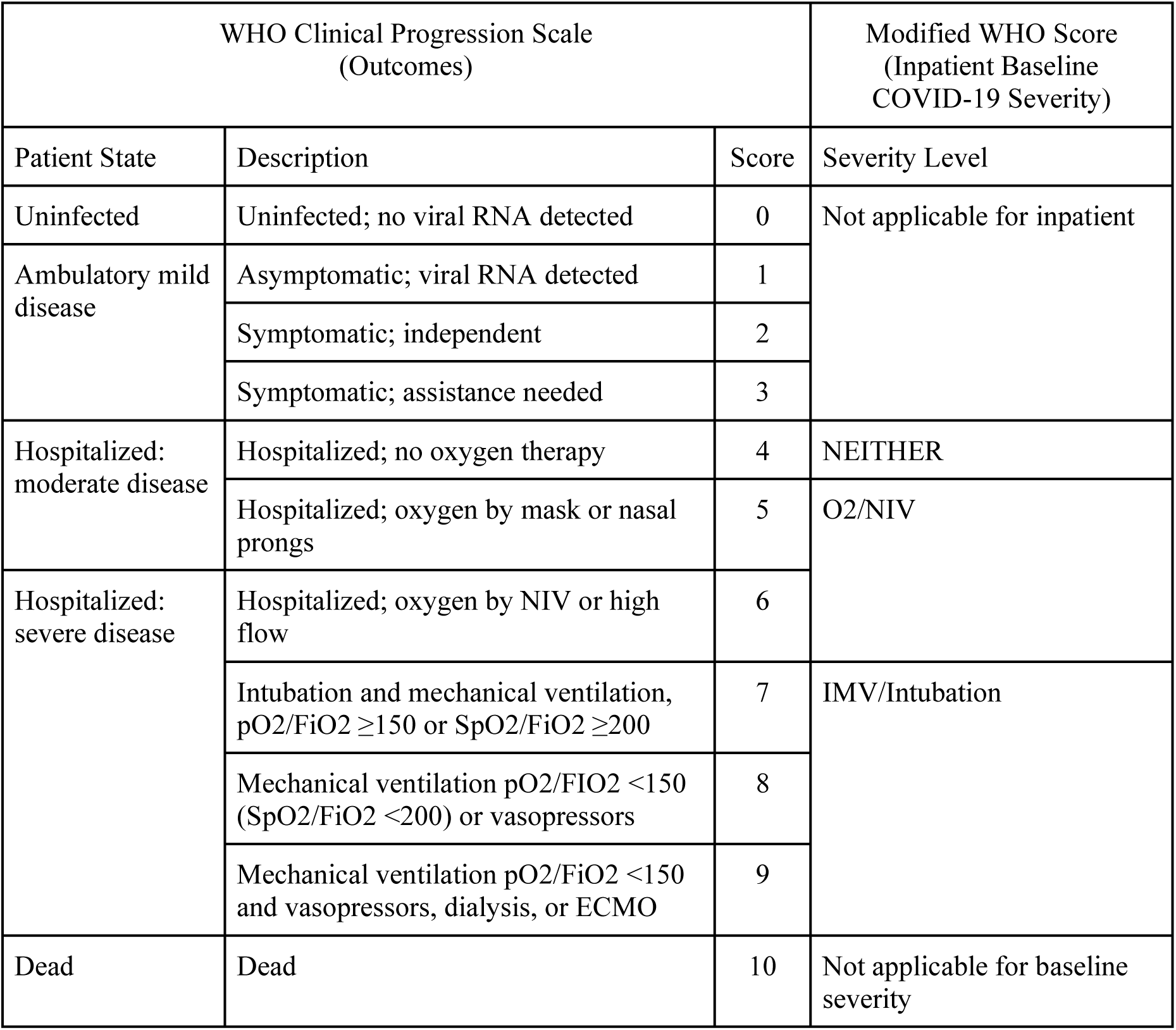
Comparison of WHO clinical progression scale to modified WHO score.

| WHO Clinical Progression Scale<br>(Outcomes) |  |  | Modified WHO Score<br>(Inpatient Baseline<br>COVID-19 Severity) |
| --- | --- | --- | --- |
| Patient State | Description | Score | Severity Level |
| Uninfected | Uninfected; no viral RNA detected | 0 | Not applicable for inpatient |
| Ambulatory mild<br>disease | Asymptomatic; viral RNA detected | 1 |  |
|  | Symptomatic; independent | 2 |  |
|  | Symptomatic; assistance needed | 3 |  |
| Hospitalized:<br>moderate disease | Hospitalized; no oxygen therapy | 4 | NEITHER |
|  | Hospitalized; oxygen by mask or nasal<br>prongs | 5 | O2/NIV |
| Hospitalized:<br>severe disease | Hospitalized; oxygen by NIV or high<br>flow | 6 |  |
| | Intubation and mechanical ventilation,<br>$pO_2/FiO_2 \geq 150$ or $SpO_2/FiO_2 \geq 200$ | 7 | IMV/Intubation |
| | Mechanical ventilation $pO_2/FiO_2 < 150$<br>($SpO_2/FiO_2 < 200$) or vasopressors | 8 | |
| | Mechanical ventilation $pO_2/FiO_2 < 150$<br>and vasopressors, dialysis, or ECMO | 9 | |
| Dead | Dead | 10 | Not applicable for baseline<br>severity |

To operationalize the score, patients were assigned to mWHO categories based on the highest severity recorded on the treatment index date, considering both the first day of treatment and the day prior to minimize potential misclassification from situations in which patients received O2 or IMV in other medical settings (e.g., emergency room or ambulance), or cases where the billing date for a procedure was captured at a later calendar date than the procedure was performed due to a lag in reporting.

In each data source, two different algorithms were developed to identify the COVID-19 severity levels (see summary in Table 2). First, we developed procedure-based algorithms that were similar for both data sources (*initial-EHR* and *initial-administrative*), except procedure-based charge codes were only applicable for the administrative chargemaster data. Next, to increase the specificity of O2 and IMV identification, we expanded the algorithms to include diagnoses indicating a clinical need for O2/NIV (hypoxia or hypoxemia) or IMV (acute respiratory distress syndrome), among other additions. The *expanded-EHR* algorithm included available clinical patient vitals and day-level diagnosis data. We then applied the EHR-based learnings to the *expanded-administrative* algorithm, adapting as needed to analogous data available in the administrative dataset. For example, day-level diagnosis data was not available in the administrative data during the hospitalizations, but we were able to utilize the flag indicating ‘present at admission’ to differentiate the admitting diagnoses from all diagnoses reported at discharge.

**Table 2.** Algorithm measure definitions to identify the COVID-19 severity score.

|  |  | Algorithm-Dataset |  |  |  |
| --- | --- | --- | --- | --- | --- |
|  |  | Initial-EHR | Initial-Administrative | Expanded-EHR | Expanded-Administrative |
| **IMV/**<br>**Intubation** | **Any of the following:** |  |  |  |  |
|  | Procedure codes (HCPCS/CPT, ICD-10-PCS) for IMV, intubation, or ECMO | ✓ | ✓ | ✓ | ✓ |
|  | Diagnosis code (ICD-10-CM)* indicating a procedure for intubation (Failed or difficult intubation) | ✓ | ✓ | ✓ | ✓ |
|  | Chargemaster Charge Code indicating procedure for IMV, intubation, or ECMO | NA | ✓ | NA | ✓ |
|  | Diagnosis code (ICD-10-CM) indicating clinical need for an IMV, intubation, or ECMO procedure (acute respiratory distress syndrome) |  |  | ✓ | ✓ |
| **O2/NIV** | **Not qualifying for IMV/intubation and any of the following:** |  |  |  |  |
|  | Procedure codes (HCPCS/CPT, ICD-10-PCS) for O2, NIV, or ventilation NOS† | ✓ | ✓ | ✓ | ✓ |
|  | Diagnosis code (ICD-10-CM)* indicating a procedure for O2 or ventilation NOS† (Dependence on supplemental oxygen) | ✓ | ✓ | ✓ | ✓ |
|  | Chargemaster Charge Code indicating procedure for O2 or NIV | NA | ✓ | NA | ✓ |
|  | Diagnosis code (ICD-10-CM) indicating clinical need for procedure (hypoxia, hypoxemia) |  |  | ✓ | ✓ |
|  | Revenue Code indicating O2 |  |  | ✓ | ✓ |
|  | Patient vitals indicating low O2 saturation (SpO2<94%), high respiratory rate (>30), or low ratio of arterial oxygen partial pressure to fractional inspired oxygen (PaO2/FiO2<300) |  |  | ✓ | U |
|  | Departmental Code indicating O2 |  |  | U | ✓ |
| **NEITHER** | **Not qualifying for IMV/intubation nor O2/NIV** |  |  |  |  |

ECMO, extracorporeal membrane oxygenation; EHR, electronic health records; IMV, invasive mechanical ventilation; NA, not applicable; O2, supplemental oxygen; NIV, non-invasive ventilation; U, unavailable in the data
\*Chargemaster encounters where day-level diagnoses are not available, diagnosis was based on present at admission only.
†When there was a ventilation procedure without invasive specification (ventilation NOS) that started or continued immediately after a procedure or diagnosis indicating an intubation or IMV event, we assumed the remaining duration of the ventilation event to be IMV. Otherwise, it was assumed to be NIV.

### 2.3. Study population

In each dataset, we identified patients hospitalized with confirmed COVID-19 (diagnosis or positive SARS-CoV-2 laboratory results) between March and August 2020 with at least 1 encounter during the 183-day baseline and no prior CSI use during the 90-day washout period. From these populations, we selected primary cohorts of DEX initiators (DEX+) and a random selection of patients who had not or had not yet initiated any CSI (CSI-) matched 1:1 on date of DEX initiation, age, sex, Charlson-Quan comorbidity score over the 183-day baseline period ***{Quan 2005}***, days since admission, and the mWHO severity level (NEITHER, O2/NIV, IMV) on treatment index (see NCT04926571 protocol for additional detail). The primary analysis cohorts matched on mWHO severity using the initial algorithms, while secondary analysis cohorts were rematched using the expanded algorithms.

### 2.4. Approach / statistical analysis

The primary analysis evaluated the treatment-specific distributions of patients in each mWHO severity level, according to both the initial algorithms that patients were matched on and the expanded algorithms. This was done to describe how many of the patients from each initial level would have shifted to a higher severity level via the expanded algorithms (the percent of NEITHER who shifted to O2/NIV or IMV, and the percent of O2/NIV that shifted to IMV). We started with the EHR data and then repeated the same process using the administrative data.

We performed two secondary analyses and one sensitivity analysis largely focusing on the administrative data. The first secondary analysis rematched cohorts in both datasets using the expanded algorithms to illustrate the distribution of patients who qualified for O2/NIV and IMV via each measure component (i.e., procedures, medical diagnosis claims, chargemaster diagnosis data, and revenue codes) using Venn diagrams to understand the relative contribution of each component type. The second compared the distribution of select patient characteristics in each treatment arm of the administrative data cohort in the categories they were matched on using the *initial-administrative* algorithm to the same cohort recategorized using the *expanded-administrative* algorithm. Lastly, a sensitivity analysis evaluated the shifts in severity and the relative contribution of each component type among a subset of patients in the administrative data cohort who were early initiators (initiated treatment on the same day or 1 day after the admission date).

All analyses were conducted using the Aetion Evidence Platform® (2021), a software for real-world data analysis, validated for a range of studies. ***{Wang 2016}*** Visualizations were created using the *tidyverse* ***{Wickham 2020}*** and *plotly* ***{Sievert 2019}*** packages in R (v4.0.3).

## 3. RESULTS

### 3.1. Primary analyses

In the EHR-based cohort with 1,768 DEX+ and CSI− patient pairs matched using the *initial-HER* algorithm, 17% were categorized as O2/NIV, 2% as IMV, and 81% as NEITHER (**Figure 1**). Among those categorized as NEITHER, use of the *expanded-EHR* algorithm shifted 56% DEX+ and 32% CSI- to O2/NIV, and 5% DEX+ and 3% CSI- to IMV. Among the patients initially categorized as O2/NIV, 10% DEX+ and 8% CSI-patients shifted to IMV.

**Figure 1.**
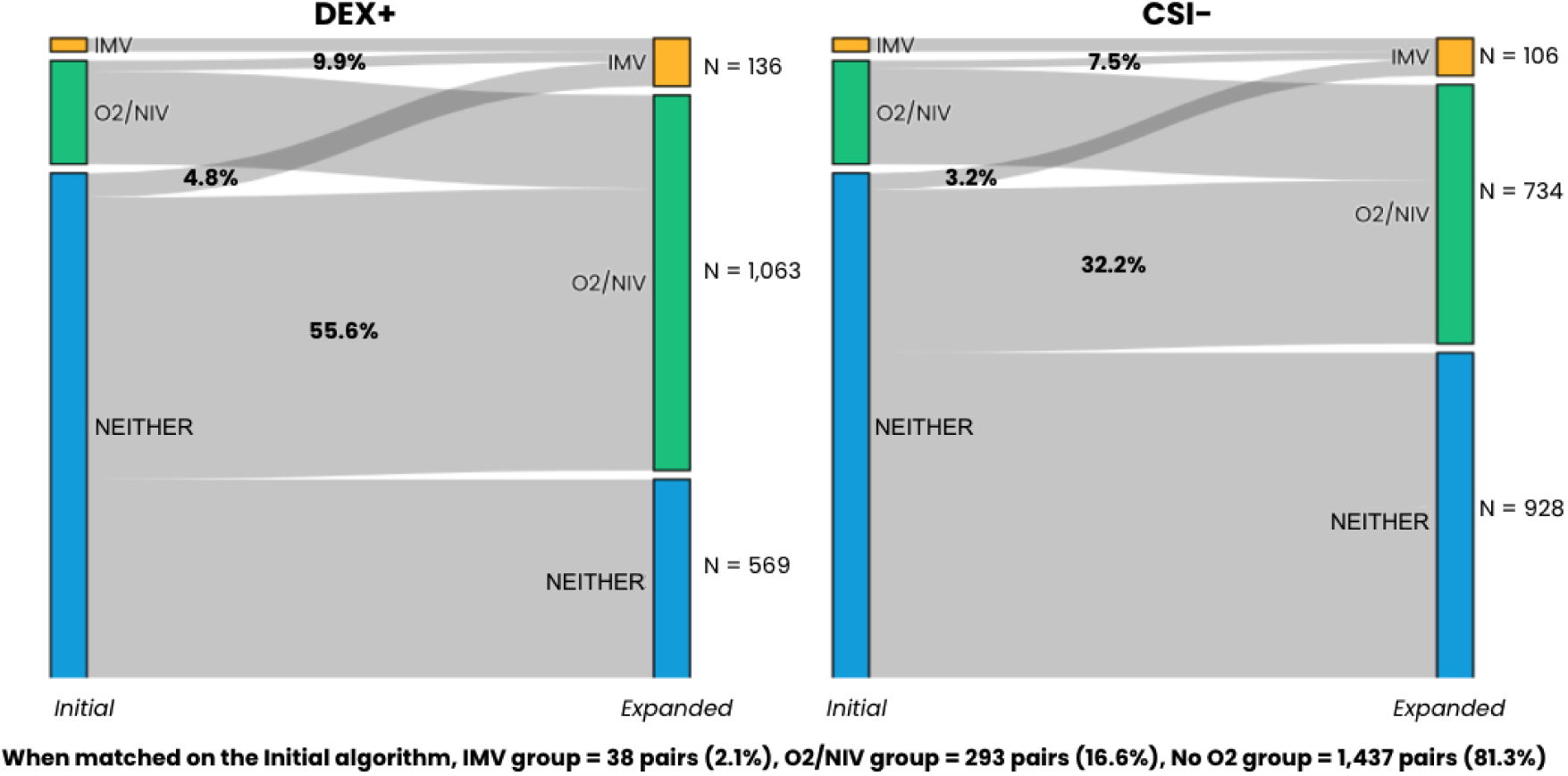
Shifts in COVID-19 severity score in each treatment arm in the EHR-based cohort matched using the initial-EHR algorithm (n=3,536; 1,768 pairs)

In the administrative data cohort with 5,524 DEX+ and CSI− patient pairs matched using the *initial-administrative* algorithm, 30% were categorized as O2/NIV, 5% as IMV, and 65% as NEITHER (**Figure 2**). Similar to the EHR-based cohort, among those categorized as NEITHER, use of the *expanded-administrative* algorithm shifted 54% DEX+ and 28% CSI- to O2/NIV, and 2% DEX+ and 1% CSI- to IMV. Among the patients initially categorized as O2/NIV, 7% DEX+ and 3% CSI− patients shifted to IMV.

**Figure 2.**
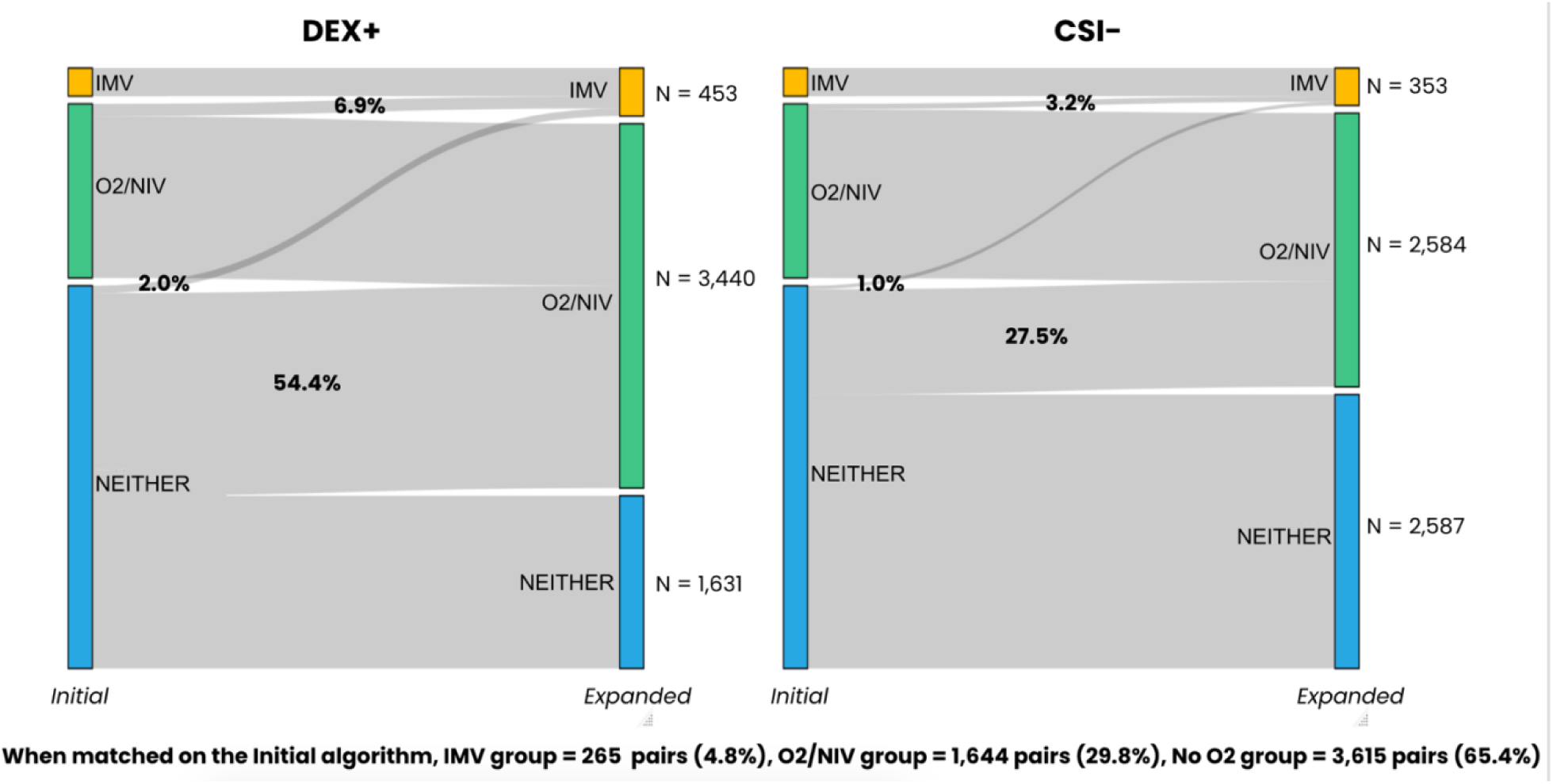
Shifts in COVID-19 severity score in each treatment arm in the administrative data cohort matched using the initial-administrative algorithm (n=11,048; 5,524 pairs)

### 3.2. Secondary and sensitivity analyses

After matching using the *expanded-administrative* algorithm, the sample size slightly decreased to 5,337 DEX+ and CSI− patient pairs with 61% categorized as O2/NIV, 7% as IMV, and 32% as NEITHER (Appendix **Figure B.1**; Appendix **Table B.1**). Illustrated areas of overlap indicate the distribution of patients with more than one measure component that qualifies them for either O2/NIV or IMV, while areas of non-overlap indicate only one qualifying component. Among the patients categorized as O2/NIV, 49% of both DEX+ and CSI− qualified only due to the added clinical diagnosis of hypoxia or hypoxemia of the *expanded-administrative* algorithm (i.e., did not have a procedure-based component). In contrast, 27% of DEX+ and 24% of CSI− qualified as IMV due to only the clinical diagnosis of acute respiratory distress syndrome. The inclusion of revenue codes only added 1 DEX+ patient to the O2/NIV group.

Additional analyses of the EHR-based cohort matched using the *expanded-EHR* algorithm are shown in Appendix B. The addition of clinical diagnoses in this expanded algorithm had a similar impact on the mWHO severity categorization to that seen in the *expanded-administrative* algorithm. Additionally, patient vitals were responsible for 14% of DEX+ and 27% of CSI− patients categorized as O2/NIV (Appendix **Figure B.2**).

Compared to the mWHO severity categorization using the *initial-administrative* algorithm that the patients were matched on, recategorization using the *expanded-administrative* algorithm showed that there was a significant decrease in the IMV group for heart disease for DEX+ patients (ASD: 0.12; **Table 3**). There was a treatment-differential change that was greater for DEX+ versus CSI− patients in the NEITHER, O2/NIV, and IMV groups for diabetes (ASD: 0.04 *vs* 0.02, 0.05 *vs* 0.02, and 0.08 *vs* 0.06), NEITHER and O2/NIV groups for age (ASD: 0.08 *vs* 0.07 and 0.04 *vs* 0.02), and in the IMV group for heart disease (ASD: 0.12 *vs* 0.01) chronic pulmonary disease (ASD: 0.09 *vs* 0.04, and end-stage renal disease (ASD: 0.09 *vs* 0.07). There was also a treatment-differential change that was less for DEX+ versus CSI− patients in the IMV group for age (ASD: 0.04 *vs* 0.05).

**Table 3.** Comparison of the distribution of select patient characteristics in each treatment arm in the administrative data cohort matched using the initial-administrative algorithm before and after recategorization using the expanded-administrative algorithm (n=11,048; 5,524 pairs)

|  | DEX+ |  |  | CSI- |  |  |
| --- | --- | --- | --- | --- | --- | --- |
|  | Matched Initial-administrative Algorithm (n=5,524) | Recategorized Expanded-administrative Algorithm (n=5,524) | ASD* | Matched Initial-administrative Algorithm (n=5,524) | Recategorized Expanded-administrative Algorithm (n=5,524) | ASD* |
|  | **NEITHER** |  |  |  |  |  |
| **N** | 3,615 | 1,631 |  | 3,615 | 2,587 |  |
| **Percent of total** | 65.4% | 29.5% |  | 65.4% | 46.8% |  |
| Age, mean (SD) | 62.18 (18.58) | 60.68 (19.77) | 0.08 | 62.14 (18.63) | 60.79 (19.34) | 0.07 |
| Male Sex, % | 48.2% | 45.2% | 0.06 | 48.2% | 47.3% | 0.02 |
| **Select comorbidities over the 183-day baseline period** |  |  |  |  |  |  |
| Diabetes, % | 38.3% | 36.3% | 0.04 | 36.9% | 35.8% | 0.02 |
| Heart Disease, % | 13.9% | 13.7% | 0.01 | 16.8% | 17.1% | 0.01 |
| Chronic Pulmonary Disease, % | 13.7% | 13.3% | 0.01 | 15.4% | 14.9% | 0.01 |
| Severe Liver Disease, % | 0.7% | 0.9% | 0.02 | 1.5% | 1.9% | 0.03 |
| End-Stage Renal Disease, % | 3.7% | 3.7% | < 0.01 | 4.0% | 3.7% | 0.02 |
|  | **O2/NIV** |  |  |  |  |  |
| **N** | 1,644 | 3,440 |  | 1,644 | 2,584 |  |
| **Percent of total** | 29.8% | 62.3% |  | 29.8% | 46.8% |  |
| Age, mean (SD) | 64.76 (16.62) | 64.06 (17.16) | 0.04 | 64.75 (16.61) | 65.14 (16.47) | 0.02 |
| Male Sex, % | 51.1% | 50.4% | 0.01 | 51.1% | 50.9% | < 0.01 |
| **Select comorbidities over the 183-day baseline period** |  |  |  |  |  |  |
| Diabetes, % | 44.5% | 42.0% | 0.05 | 42.3% | 41.3% | 0.02 |
| Heart Disease, % | 16.4% | 15.3% | 0.03 | 19.4% | 18.2% | 0.03 |
| Chronic Pulmonary Disease, % | 16.9% | 15.4% | 0.04 | 20.3% | 19.0% | 0.03 |
| Severe Liver Disease, % | 0.8% | 0.7% | 0.01 | 1.2% | 1.0% | 0.02 |
| End-Stage Renal Disease, % | 3.3% | 3.7% | 0.02 | 5.2% | 5.1% | < 0.01 |
|  | **IMV** |  |  |  |  |  |
| **N** | 265 | 453 |  | 265 | 353 |  |
| **Percent of total** | 4.8% | 8.2% |  | 4.8% | 6.4% |  |
| Age, mean (SD) | 64.62 (12.82) | 64.11 (14.08) | 0.04 | 64.88 (12.65) | 64.27 (13.60) | 0.05 |
| Male Sex, % | 62.6% | 61.1% | 0.03 | 62.6% | 60.1% | 0.05 |
| **Select comorbidities over the 183-day baseline period** |  |  |  |  |  |  |
| Diabetes, % | 52.8% | 48.8% | 0.08 | 54.7% | 51.6% | 0.06 |
| Heart Disease, % | 22.6% | 17.9% | **0.12** | 18.9% | 18.7% | 0.01 |
| Chronic Pulmonary Disease, % | 23.0% | 19.2% | 0.09 | 20.4% | 19.0% | 0.04 |
| Severe Liver Disease, % | 1.1% | 0.9% | 0.02 | 2.6% | 2.3% | 0.02 |
| End-Stage Renal Disease, % | 5.7% | 3.8% | 0.09 | 8.7% | 6.8% | 0.07 |
| *ASD, Absolute Standardized Difference to compare the expanded algorithm to the initial algorithm (bold text indicates ASD>0.1 considered to be a significant difference) |  |  |  |  |  |  |

When the administrative cohort was restricted to early initiators in the sensitivity analysis, similar shifts in mWHO severity and relative contributions of each component type were identified (**Figures B.3-B.4**).

## 4. CONCLUSION

Application of the expanded algorithms resulted in treatment-differential shifts to higher mWHO severity levels in both datasets, suggesting a substantial increase in the specificity. As expected, IMV was mostly captured via procedure-related encounters, but there were still notable shifts from NEITHER and from O2/NIV to IMV. In contrast, the shifts from NEITHER to O2/NIV under the expanded algorithms were substantial. More than half of the DEX+ patients (56% and 49% in the EHR and administrative data cohorts, respectively) and more than one-quarter of the CSI− patients (32% and 28%) had no evidence of an O2/NIV procedure-related encounter, but had one of the other O2/NIV components (largely diagnosis of hypoxia or hypoxemia). Rematching the administrative data cohort using the *expanded-administrative* algorithm further supported the value of adding the diagnoses indicating clinical need for procedure. There were important treatment-differential changes in the distributions of patient characteristics seen when patients who had been matched using the *initial-administrative* algorithm had been recategorized via the *expanded-administrative* algorithm that would have been masked had a procedure-based algorithm alone been used.

Since the patients randomized in the RECOVERY trial were older and had more comorbidities ***{RECOVERY 2020}*** than our real-world administrative data cohort, we anticipated that our patients may be less severe at baseline. However, we were surprised to see that compared to the RECOVERY trial DEX+ treatment arm, the *initial-administrative* algorithm identified 42% more patients in the NEITHER group, and 31% and 11% less patients in the O2/NIV and IMV groups, respectively. After matching using the *expanded-administrative* algorithm, the severity classifications aligned with our expectations, i.e., we had 8% more patients in the NEITHER group compared with RECOVERY, approximately the same in O2/NIV, and 8% less patients in IMV.

Advantages and limitations of the HealthVerity administrative data were taken into consideration. First, although the included open claims data has the benefit of near-real-time capture, it may be less complete for the most recent calendar dates. However, because our study period ended August 2020 to align with the Optum data we had available, this concern was minimal. Additionally, although access to inpatient medication use via the chargemaster data is a benefit over most other RWD sources, inpatient corticosteroids (DEX and other CSIs) are captured via non-standardized vendor descriptions rather than standardized fields (e.g., national drug codes). We have therefore developed a series of search string algorithms in collaboration with a licensed pharmacist to minimize the potential for misclassification of these medications. Finally, while inpatient day-level diagnoses are not consistently available in HealthVerity, which could lead to under-reporting of severity on days during the hospitalization, we were able to incorporate the admitting diagnoses available via the chargemaster data into the *expanded-administrative algorithm* and we assume to have complete diagnosis data for the majority of patients since 90% of our cohort were early initiators with a treatment index either on or one day after the admission date. This concern was further mitigated by the finding that the results among early initiators were similar.

Access to the Optum data with patient vitals and day-level diagnoses informed and improved our administrative data algorithm. The fact that the shifts using the administrative data cohort were similar to the shifts in the EHR-based cohort provides additional assurance that we were able to still capture necessary diagnoses via admitting diagnoses alone. Therefore, the expanded algorithms have applicability for use in any RWD that allows for valid identification of inpatient hospitalization and includes procedure codes. While day-level diagnoses are ideal, access to admitting diagnoses should be sufficient (in particular when study entry happens close to admission) with noted limitations.

Applying learnings from use of an EHR-based algorithm to our administrative data algorithm minimized treatment-differential misclassification, and allowed for more specific identification of lower and higher severity patients through the addition of relevant clinical diagnoses as well as patient vitals (where available). These findings demonstrate the importance of these expanded algorithms, as O2 and NIV, which are critical to COVID-19 severity categorization, may be substantially under-reported in RWD using procedure-based encounters alone. Although further validation efforts may be considered, our expanded algorithms are an important addition to the COVID-19 RWD literature with applicability for any study requiring categorization of disease severity in inpatient EHR or administrative data.

## Data Availability

Patient-level data belong to third parties and cannot be shared publicly. Upon reasonable request, researchers may obtain limited access to data and analytics infrastructure.

## Acknowledgements

We wish to thank the following additional members of the COVID-19 Research Collaborative team for their additional review and discussion of the research design and interpretation of findings: Marie C. Bradley, PhD, MScPH, MPharm; Silvia Perez-Vilar, PharmD, PhD; Laura M. Roe, MMCi (U.S. Food and Drug Administration), Amy Abernethy, MD, PhD (former Principal Deputy Commissioner, U.S. Food & Drug Administration), Nevine Zariffa, M.Math. (NMD Group LLC consulting), and Anthony Louder, PhD (Aetion, Inc.). We wish to also thank Melanie Wang, MPH (Aetion, Inc.) for her management and coordination of this collaborative research effort.

## ABBREVIATIONS

COVID-19: coronavirus disease 19
CPT: current procedural terminology
CSI: corticosteroid of interest (DEX, methylprednisolone, prednisone, hydrocortisone)
CSI-: random selection of patients who had not (yet) initiated any CSI
DEX: dexamethasone
DEX+: dexamethasone initiators in the matched cohort
ECMO: extracorporeal membrane oxygenation
EHR: electronic health record
FDA: United States Food and Drug Administration
HCPCS: healthcare common procedure coding system
ICD-10-CM: international classification of disease codes, 10th revision, clinical modification
ICD-10-PCS: international classification of disease codes, 10th revision, procedural classification system
IMV: invasive mechanical ventilation
mWHO: modified version of WHO Clinical Progression Scale for COVID-19 severity
NEITHER: neither O2/NIV nor IMV
NIV: noninvasive ventilation
O2: oxygen/supplemental oxygen
PaO2/FIO2: ratio of arterial oxygen partial pressure to fractional inspired oxygen
RWD: real-world data
SpO2: blood oxygen saturation
US: United States
WHO: World Health Organization

## APPENDIX A: mWHO COVID-19 Severity Detailed Breakdown and Code Lists

**Table A.1.** Overview of the expanded-administrative algorithm used in HealthVerity data.

| COVID-19 Severity | Algorithm |
| --- | --- |
| IMV | IP Chargemaster, OP Chargemaster, or Medical Claims with procedure codes (*HCPCS/CPT, ICD-10-PCS*) for **Intubation, IMV, or ECMO** |
|  | IP Chargemaster or OP Chargemaster with *Charge Code* matches for:<br>(any terms in the **Intubation Inclusion** search term list **AND** does **NOT** match any terms in the **Intubation/Ventilation Exclusion** search term list);<br>OR<br>(any terms in the **Ventilation Inclusion** search term list **AND** matches <u>at least one</u> term in the **IMV Inclusion** search term list **AND** does **NOT** match any terms in the **Intubation/Ventilation Exclusion** term list **AND** does **NOT** match any terms in the **IMV Exclusion** term list);<br>OR<br>(any terms in the **ECMO Inclusion** search term list **AND** does **NOT** match any terms in the **ECMO Exclusion** search term list) |
|  | IP Chargemaster*, OP Chargemaster*, or Medical Claims with a diagnosis code (*ICD-10-CM*) indicating a procedure or clinical need for **intubation** or **IMV** (acute respiratory distress syndrome) |
| O2/NIV | IP Chargemaster, OP Chargemaster, or Medical Claims with procedure codes (*HCPCS/CPT, ICD-10-PCS*) for **NIV** or **ventilation not otherwise specified as invasive (NOS)** or **Supplemental O2** |
|  | IP Chargemaster or OP Chargemaster with *Charge Code* matches for<br>(any terms in the **Ventilation Inclusion** search term list **AND** matches <u>at least one</u> term in the **NIV Inclusion** search term list **AND** does **NOT** match any terms in the **Intubation/Ventilation Exclusion** search term list);<br>OR<br>(any terms in the **Ventilation Inclusion** search term list **AND** does **NOT** match any terms in the **Intubation/Ventilation Exclusion** search term list **AND** does **NOT** match any terms in the **IMV Inclusion** term list **AND** does **NOT** match any terms in the **NIV Inclusion** term list);<br>OR<br>(any term in the **Oxygen Inclusion** search term list **AND** does **NOT** match any terms in the **Oxygen Exclusion** search term list) |
|  | IP Chargemaster*, OP Chargemaster*, or Medical Claims with a diagnosis code (*ICD-10-CM*) indicating a procedure or clinical need for **ventilation NOS** or **Supplemental O2** (hypoxia, hypoxemia) |
|  | Medical Claims with a *Revenue Code* for **Supplemental O2** |
|  | IP Chargemaster or OP Chargemaster with a *Department Code* or *Standard Department Code* with the **Supplemental O2** search term |

\*Diagnosis code in admitting position only (i.e., when 'present at admission' flag is true)

**Table A.2.** Code list for the expanded-administrative algorithm.

| Component | Measure Detail | Type | Code | Description |
| --- | --- | --- | --- | --- |
| Intubation | Procedure code | HCPCS/CPT | 31500 | Intubation, endotracheal, emergency procedure |
|  |  | ICD-10-PCS | 09HN7BZ | Insertion of Airway into Nasopharynx, Via Natural or Artificial Opening |
|  |  | ICD-10-PCS | 0DH57BZ | Insertion of Airway into Esophagus, Via Natural or Artificial Opening |
|  |  | ICD-10-PCS | 0DH58BZ | Insertion of Airway into Esophagus, Via Natural or Artificial Opening Endoscopic |
|  |  | ICD-10-PCS | 0WHQ73Z | Insertion of Infusion Device into Respiratory Tract, Via Natural or Artificial Opening |
|  |  | ICD-10-PCS | 0WHQ7YZ | Insertion of Other Device into Respiratory Tract, Via Natural or Artificial Opening |
|  |  | ICD-10-PCS | 09HN8BZ | Insertion of Airway into Nasopharynx, Via Natural or Artificial Opening Endoscopic |
|  |  | ICD-10-PCS | 0B21XEZ | Change Endotracheal Airway in Trachea, External Approach |
|  |  | ICD-10-PCS | 0BH13EZ | Insertion of Endotracheal Airway into Trachea, Percutaneous Approach |
|  |  | ICD-10-PCS | 0BH17EZ | Insertion of Endotracheal Airway into Trachea, Via Natural or Artificial Opening |
|  |  | ICD-10-PCS | 0BH18EZ | Insertion of Endotracheal Airway into Trachea, Via Natural or Artificial Opening Endoscopic |
|  |  | ICD-10-PCS | 0CHY7BZ | Insertion of Airway into Mouth and Throat, Via Natural or Artificial Opening |
|  |  | ICD-10-PCS | 0CHY8BZ | Insertion of Airway into Mouth and Throat, Via Natural or Artificial Opening Endoscopic |
|  | Dx indicating procedure | ICD-10-CM | T88.4 | Failed or difficult intubation |
|  |  | ICD-10-CM | T88.4XXA | Failed or difficult intubation, initial encounter |
|  |  | ICD-10-CM | T88.4XXD | Failed or difficult intubation, subsequent encounter |
|  |  | ICD-10-CM | T88.4XXS | Failed or difficult intubation, sequela |
| IMV | Procedure code | HCPCS/CPT | E0472 | Respiratory assist device, bi-level pressure capability, with backup rate feature, used with invasive interface, e.g., tracheostomy tube (intermittent assist device with continuous positive airway pressure device) |
|  |  | HCPCS/CPT | K0534 | Respiratory assist device, bi-level pressure capability, with backup rate feature, used with invasive interface, e.g., tracheostomy tube (intermittent assist device with continuous positive airway pressure device) |
|  | Dx indicating clinical need for procedure | ICD-10-CM | J80 | Acute respiratory distress syndrome |
| ECMO | Procedure code | HCPCS/CPT | 33946 | Extracorporeal membrane oxygenation (ECMO)/extracorporeal life support (ECLS) provided by physician; initiation, veno-venous |
|  |  | HCPCS/CPT | 33947 | Extracorporeal membrane oxygenation (ECMO)/extracorporeal life support (ECLS) provided by physician; initiation, veno-arterial |
|  |  | HCPCS/CPT | 33948 | Extracorporeal membrane oxygenation (ECMO)/extracorporeal life support (ECLS) provided by physician; daily management, each day, veno-venous |
|  |  | HCPCS/CPT | 33949 | Extracorporeal membrane oxygenation (ECMO)/extracorporeal life support (ECLS) provided by physician; daily management, each day, veno-arterial |
|  |  | HCPCS/CPT | 33951 | Extracorporeal membrane oxygenation (ECMO)/extracorporeal life support (ECLS) provided by physician; insertion of peripheral (arterial and/or venous) cannula(e), percutaneous, birth through 5 years of age (includes fluoroscopic guidance, when performed) |
| ECMO | Procedure code | HCPCS/CPT | 33952 | Extracorporeal membrane oxygenation (ECMO)/extracorporeal life support (ECLS) provided by physician; insertion of peripheral (arterial and/or venous) cannula(e), percutaneous, 6 years and older (includes fluoroscopic guidance, when performed) |
|  |  | HCPCS/CPT | 33953 | Extracorporeal membrane oxygenation (ECMO)/extracorporeal life support (ECLS) provided by physician; insertion of peripheral (arterial and/or venous) cannula(e), open, birth through 5 years of age |
|  |  | HCPCS/CPT | 33954 | Extracorporeal membrane oxygenation (ECMO)/extracorporeal life support (ECLS) provided by physician; insertion of peripheral (arterial and/or venous) cannula(e), open, 6 years and older |
|  |  | HCPCS/CPT | 33955 | Extracorporeal membrane oxygenation (ECMO)/extracorporeal life support (ECLS) provided by physician; insertion of central cannula(e) by sternotomy or thoracotomy, birth through 5 years of age |
|  |  | HCPCS/CPT | 33956 | Extracorporeal membrane oxygenation (ECMO)/extracorporeal life support (ECLS) provided by physician; insertion of central cannula(e) by sternotomy or thoracotomy, 6 years and older |
|  |  | HCPCS/CPT | 33957 | Extracorporeal membrane oxygenation (ECMO)/extracorporeal life support (ECLS) provided by physician; reposition peripheral (arterial and/or venous) cannula(e), percutaneous, birth through 5 years of age (includes fluoroscopic guidance, when performed) |
|  |  | HCPCS/CPT | 33958 | Extracorporeal membrane oxygenation (ECMO)/extracorporeal life support (ECLS) provided by physician; reposition peripheral (arterial and/or venous) cannula(e), percutaneous, 6 years and older (includes fluoroscopic guidance, when performed) |
|  |  | HCPCS/CPT | 33959 | Extracorporeal membrane oxygenation (ECMO)/extracorporeal life support (ECLS) provided by physician; reposition peripheral (arterial and/or venous) cannula(e), open, birth through 5 years of age (includes fluoroscopic guidance, when performed) |
|  |  | HCPCS/CPT | 33962 | Extracorporeal membrane oxygenation (ECMO)/extracorporeal life support (ECLS) provided by physician; reposition peripheral (arterial and/or venous) cannula(e), open, 6 years and older (includes fluoroscopic guidance, when performed) |
|  |  | HCPCS/CPT | 33963 | Extracorporeal membrane oxygenation (ECMO)/extracorporeal life support (ECLS) provided by physician; reposition of central cannula(e) by sternotomy or thoracotomy, birth through 5 years of age (includes fluoroscopic guidance, when performed) |
|  |  | HCPCS/CPT | 33964 | Extracorporeal membrane oxygenation (ECMO)/extracorporeal life support (ECLS) provided by physician; reposition central cannula(e) by sternotomy or thoracotomy, 6 years and older (includes fluoroscopic guidance, when performed) |
|  |  | HCPCS/CPT | 33965 | Extracorporeal membrane oxygenation (ECMO)/extracorporeal life support (ECLS) provided by physician; removal of peripheral (arterial and/or venous) cannula(e), percutaneous, birth through 5 years of age |
|  |  | HCPCS/CPT | 33966 | Extracorporeal membrane oxygenation (ECMO)/extracorporeal life support (ECLS) provided by physician; removal of peripheral (arterial and/or venous) cannula(e), percutaneous, 6 years and older |
|  |  | HCPCS/CPT | 33969 | Extracorporeal membrane oxygenation (ECMO)/extracorporeal life support (ECLS) provided by physician; removal of peripheral (arterial and/or venous) cannula(e), open, birth through 5 years of age |
|  |  | HCPCS/CPT | 33984 | Extracorporeal membrane oxygenation (ECMO)/extracorporeal life support (ECLS) provided by physician; removal of peripheral (arterial and/or venous) cannula(e), open, 6 years and older |
|  |  | HCPCS/CPT | 33985 | Extracorporeal membrane oxygenation (ECMO)/extracorporeal life support (ECLS) provided by physician; removal of central cannula(e) by sternotomy or thoracotomy, birth through 5 years of age |
|  |  | HCPCS/CPT | 33986 | Extracorporeal membrane oxygenation (ECMO)/extracorporeal life support (ECLS) provided by physician; removal of central cannula(e) by sternotomy or thoracotomy, 6 years and older |
|  |  | HCPCS/CPT | 33987 | Arterial exposure with creation of graft conduit (eg, chimney graft) to facilitate arterial perfusion for ECMO/ECLS (List separately in addition to code for primary procedure) |
| ECMO | Procedure code | HCPCS/CPT | 33988 | Insertion of left heart vent by thoracic incision (eg, sternotomy, thoracotomy) for ECMO/ECLS |
|  |  | HCPCS/CPT | 33989 | Removal of left heart vent by thoracic incision (eg, sternotomy, thoracotomy) for ECMO/ECLS |
|  |  | HCPCS/CPT | 36822 | Insertion of cannula(s) for prolonged extracorporeal circulation for cardiopulmonary insufficiency (ECMO) (separate procedure) |
|  |  | HCPCS/CPT | J0170 | INJECTION, ADRENALIN, EPINEPHRINE, UP TO 1 ML AMPULE |
|  |  | HCPCS/CPT | J0171 | INJECTION, ADRENALIN, EPINEPHRINE, 0.1 MG |
|  |  | HCPCS/CPT | J1265 | INJECTION, DOPAMINE HCL, 40 MG |
|  |  | HCPCS/CPT | J2370 | INJECTION, PHENYLEPHRINE HCL, UP TO 1 ML |
|  |  | HCPCS/CPT | Q4076 | INJECTION, DOPAMINE HCL, 40 MG |
|  |  | ICD-10-PCS | 5A05121 | Extracorporeal Hyperbaric Oxygenation, Intermittent |
|  |  | ICD-10-PCS | 5A05221 | Extracorporeal Hyperbaric Oxygenation, Continuous |
|  |  | ICD-10-PCS | 5A15223 | Extracorporeal Membrane Oxygenation, Continuous |
|  |  | ICD-10-PCS | 5A1522F | Extracorporeal Oxygenation, Membrane, Central |
|  |  | ICD-10-PCS | 5A1522G | Extracorporeal Oxygenation, Membrane, Peripheral Veno-arterial |
|  |  | ICD-10-PCS | 5A1522H | Extracorporeal Oxygenation, Membrane, Peripheral Veno-venous |
| NIV | Procedure code | HCPCS/CPT | 5A09459 | Assistance with Respiratory Ventilation, 24-96 Consecutive Hrs, Continuous Negative Airway Pressure |
|  |  | HCPCS/CPT | 5A09557 | Assistance with Respiratory Ventilation, Greater than 96 Consecutive Hrs, Continuous Positive Airway Pressure |
|  |  | HCPCS/CPT | 5A09357 | Assistance with Respiratory Ventilation, <24 Consecutive Hrs, Continuous Positive Airway Pressure |
|  |  | HCPCS/CPT | 5A09358 | Assistance with Respiratory Ventilation, <24 Consecutive Hrs, Intermittent Positive Airway Pressure |
|  |  | HCPCS/CPT | 5A09359 | Assistance with Respiratory Ventilation, <24 Consecutive Hrs, Continuous Negative Airway Pressure |
|  |  | HCPCS/CPT | 5A0935B | Assistance with Respiratory Ventilation, <24 Consecutive Hrs, Intermittent Negative Airway Pressure |
|  |  | HCPCS/CPT | 5A09457 | Assistance with Respiratory Ventilation, 24-96 Consecutive Hrs, Continuous Positive Airway Pressure |
|  |  | HCPCS/CPT | 5A09458 | Assistance with Respiratory Ventilation, 24-96 Consecutive Hrs, Intermittent Positive Airway Pressure |
|  |  | HCPCS/CPT | 5A0945B | Assistance with Respiratory Ventilation, 24-96 Consecutive Hrs, Intermittent Negative Airway Pressure |
|  |  | HCPCS/CPT | 5A09558 | Assistance with Respiratory Ventilation, Greater than 96 Consecutive Hrs, Intermittent Positive Airway Pressure |
|  |  | HCPCS/CPT | 5A09559 | Assistance with Respiratory Ventilation, Greater than 96 Consecutive Hrs, Continuous Negative Airway Pressure |
|  |  | HCPCS/CPT | 5A0955B | Assistance with Respiratory Ventilation, Greater than 96 Consecutive Hrs, Intermittent Negative Airway Pressure |
| Ventilation NOS | Procedure code | HCPCS/CPT | 94002 | Ventilation assist and management, initiation of pressure or volume preset ventilators for assisted or controlled breathing; hospital inpatient/observation, initial day |
|  |  | HCPCS/CPT | 94003 | Ventilation assist and management, initiation of pressure or volume preset ventilators for assisted or controlled breathing; hospital inpatient/observation, each subsequent day |
|  |  | HCPCS/CPT | 94004 | Ventilation assist and management, initiation of pressure or volume preset ventilators for assisted or controlled breathing; nursing facility, per day |
|  |  | HCPCS/CPT | 94656 | Ventilation assist and management, initiation of pressure or volume preset ventilators for assisted or controlled breathing; first day |
|  |  | HCPCS/CPT | 94657 | Ventilation assist and management, initiation of pressure or volume preset ventilators for assisted or controlled breathing; subsequent days |
|  |  | ICD-10-PCS | 5A0920Z | Assistance with Respiratory Filtration, Continuous |
|  |  | ICD-10-PCS | 5A0945Z | Assistance with Respiratory Ventilation, 24-96 Consecutive Hrs |
|  |  | ICD-10-PCS | 5A0955Z | Assistance with Respiratory Ventilation, Greater than 96 Consecutive Hrs |
|  |  | ICD-10-PCS | 5A19054 | Respiratory Ventilation, Single, Nonmechanical |
|  |  | ICD-10-PCS | 5A1935Z | Respiratory Ventilation, <24 Consecutive Hrs |
| Ventilation<br>NOS | Procedure code | ICD-10-PCS | 5A0935Z | Assistance with Respiratory Ventilation, <24 Consecutive Hrs |
|  |  | ICD-10-PCS | 5A09358 | Assistance with Respiratory Ventilation, <24 Consecutive Hrs, Intermittent Pos Airway Pressure |
|  |  | ICD-10-PCS | 5A0935B | Assistance with Respiratory Ventilation, <24 Consecutive hrs, Intermittent Neg Airway Pressure |
|  |  | ICD-10-PCS | 5A1945Z | Respiratory Ventilation, 24-96 Consecutive Hrs |
|  |  | ICD-10-PCS | 5A1955Z | Respiratory Ventilation, Greater than 96 Consecutive Hrs |
|  | Dx indicating<br>procedure | ICD-10-CM | J95.85 | Complication of respirator [ventilator] |
|  |  | ICD-10-CM | J95.851 | Ventilator associated pneumonia |
|  |  | ICD-10-CM | J95.859 | Other complication of respirator [ventilator] |
|  |  | ICD-10-CM | Z99.11 | Dependence on respirator [ventilator] status |
| Supplemental<br>O2 | Procedure code | HCPCS/CPT | 31730 | Transtacheal (percutaneous) introduction of needle wire dilator/stent or indwelling tube for oxygen |
|  |  | HCPCS/CPT | 99183 | Physician attendance & supervision of hyperbaric oxygen therapy, per session/Physician or other qualified health care professional attendance & supervision of hyperbaric oxygen therapy, per session |
|  |  | HCPCS/CPT | A4608 | Transtacheal oxygen catheter, each |
|  |  | HCPCS/CPT | A4615 | Cannula, nasal |
|  |  | HCPCS/CPT | A4616 | Tubing (oxygen), per foot |
|  |  | HCPCS/CPT | C1300 | Hyperbaric oxygen under pressure, full body chamber, per 30 minute interval |
|  |  | HCPCS/CPT | E0424 | Stationary compressed gaseous system, rental; includes container, contents, regulator, flowmeter, humidifier, nebulizer, cannula or mask, and tubing |
|  |  | HCPCS/CPT | E0425 | Stationary compressed gas system, purchase; includes regulator, flowmeter, humidifier, nebulizer, cannula or mask, and tubing |
|  |  | HCPCS/CPT | E0430 | Portable gaseous oxygen system, purchase; Includes regulator, flowmeter, humidifier, cannula or mask, and tubing |
|  |  | HCPCS/CPT | E0431 | Portable gaseous oxygen system, rental; Includes portable container, regulator, flowmeter, humidifier, cannula or mask, and tubing |
|  |  | HCPCS/CPT | E0433 | Portable liquid oxygen system, rental; Home liquefier used to fill portable liquid oxygen container; Includes portable containers, regulator, flowmeter, humidifier, cannula or mask and tubing, with or without supply reservoir and contents gauge |
|  |  | HCPCS/CPT | E0434 | Portable liquid oxygen system, rental; Includes portable container, supply reservoir, humidifier, flowmeter, refill adapter, contents gauge, cannula or mask, and tubing |
|  |  | HCPCS/CPT | E0435 | Portable liquid oxygen system, purchase; Includes portable container, supply reservoir, flowmeter, humidifier, contents gauge, cannula or mask, tubing, and refill adapter |
|  |  | HCPCS/CPT | E0439 | Stationary liquid oxygen system, rental; Includes container, contents, regulator, flowmeter, humidifier, nebulizer, cannula or mark, & tubing |
|  |  | HCPCS/CPT | E0440 | Stationary liquid oxygen system, purchase; Includes use of reservoir, contents indicator, regulator, flowmeter, humidifier, nebulizer, cannula or mark, and |
|  |  | HCPCS/CPT | E0441 | STATIONARY OXYGEN CONTENTS, GASEOUS, 1 MONTH'S SUPPLY = 1 UNIT |
|  |  | HCPCS/CPT | E0442 | STATIONARY OXYGEN CONTENTS, LIQUID, 1 MONTH'S SUPPLY = 1 UNIT |
|  |  | HCPCS/CPT | E0443 | PORTABLE OXYGEN CONTENTS, GASEOUS, 1 MONTH'S SUPPLY = 1 UNIT |
|  |  | HCPCS/CPT | E0444 | PORTABLE OXYGEN CONTENTS, LIQUID, 1 MONTH'S SUPPLY = 1 UNIT |
|  |  | HCPCS/CPT | E0455 | OXYGEN TENT, EXCLUDING CROUP OR PEDIATRIC TENTS |
|  |  | HCPCS/CPT | E1353 | REGULATOR |
|  |  | HCPCS/CPT | E1355 | STAND/RACK |
| Supplemental<br>O2 | Procedure code | HCPCS/CPT | E1390 | OXYGEN CONCENTRATOR, SINGLE DELIVERY PORT, CAPABLE OF DELIVERING 85 PERCENT OR GREATER OXYGEN CONCENTRATION AT THE PRESCRIBED FLOW RATE |
|  |  | HCPCS/CPT | E1391 | OXYGEN CONCENTRATOR, DUAL DELIVERY PORT, CAPABLE OF DELIVERING 85 PERCENT OR GREATER OXYGEN CONCENTRATION AT THE PRESCRIBED FLOW RATE, EACH |
|  |  | HCPCS/CPT | E1392 | PORTABLE OXYGEN CONCENTRATOR, RENTAL |
|  |  | HCPCS/CPT | E1405 | OXYGEN AND WATER VAPOR ENRICHING SYSTEM WITH HEATED DELIVERY |
|  |  | HCPCS/CPT | E1406 | OXYGEN AND WATER VAPOR ENRICHING SYSTEM WITHOUT HEATED DELIVERY |
|  |  | HCPCS/CPT | K0671 | PORTABLE OXYGEN CONCENTRATOR, RENTAL |
|  |  | HCPCS/CPT | K0738 | PORTABLE GASEOUS OXYGEN SYSTEM, RENTAL; HOME COMPRESSOR USED TO FILL PORTABLE OXYGEN CYLINDERS; INCLUDES PORTABLE CONTAINERS, REGULATOR, FLOWMETER, HUMIDIFIER, CANNULA OR MASK, AND TUBING |
|  |  | HCPCS/CPT | K0740 | REPAIR OR NONROUTINE SERVICE FOR OXYGEN EQUIPMENT REQUIRING THE SKILL OF A TECHNICIAN, LABOR COMPONENT, PER 15 MINUTES |
|  |  | HCPCS/CPT | K0741 | Portable gaseous oxygen system, rental; Includes portable container, regulator, flowmeter, humidifier, cannula or mask, and tubing, for cluster headaches |
|  |  | HCPCS/CPT | QH | OXYGEN CONSERVING DEVICE IS BEING USED WITH AN OXYGEN DELIVERY SYSTEM |
|  |  | HCPCS/CPT | S8120 | OXYGEN CONTENTS, GASEOUS, 1 UNIT EQUALS 1 CUBIC FOOT |
|  |  | HCPCS/CPT | S8121 | OXYGEN CONTENTS, LIQUID, 1 UNIT EQUALS 1 POUND |
|  |  | ICD-10-PCS | 31730 | Transtacheal (percutaneous) introduction of needle wire dilator/stent or indwelling tube for oxygen therapy |
|  |  | ICD-10-PCS | 99183 | Physician attendance and supervision of hyperbaric oxygen therapy, per session/Physician or other qualified health care professional attendance & supervision of hyperbaric oxygen therapy, per session |
|  |  | ICD-10-PCS | A4608 | TRANSTRACHEAL OXYGEN CATHETER, EACH |
|  |  | ICD-10-PCS | A4615 | CANNULA, NASAL |
|  |  | ICD-10-PCS | A4616 | TUBING (OXYGEN), PER FOOT |
|  |  | ICD-10-PCS | C1300 | HYPERBARIC OXYGEN UNDER PRESSURE, FULL BODY CHAMBER, PER 30 MINUTE INTERVAL |
|  |  | ICD-10-PCS | E0424 | Stationary compressed gaseous oxygen system, rental; Includes container, contents, regulator, flowmeter, humidifier, nebulizer, cannula or mask, and tubing |
|  |  | ICD-10-PCS | E0425 | STATIONARY COMPRESSED GAS SYSTEM, PURCHASE; INCLUDES REGULATOR, FLOWMETER, HUMIDIFIER, NEBULIZER, CANNULA OR MASK, AND TUBING |
|  |  | ICD-10-PCS | E0430 | PORTABLE GASEOUS OXYGEN SYSTEM, PURCHASE; INCLUDES REGULATOR, FLOWMETER, HUMIDIFIER, CANNULA OR MASK, AND TUBING |
|  |  | ICD-10-PCS | E0431 | PORTABLE GASEOUS OXYGEN SYSTEM, RENTAL; INCLUDES PORTABLE CONTAINER, REGULATOR, FLOWMETER, HUMIDIFIER, CANNULA OR MASK, AND TUBING |
|  |  | ICD-10-PCS | E0433 | PORTABLE LIQUID OXYGEN SYSTEM, RENTAL; HOME LIQUEFIER USED TO FILL PORTABLE LIQUID OXYGEN CONTAINERS, INCLUDES PORTABLE CONTAINERS, REGULATOR, FLOWMETER, HUMIDIFIER, CANNULA OR MASK AND TUBING, WITH OR WITHOUT SUPPLY RESERVOIR AND CONTENTS GAUGE |
|  |  | ICD-10-PCS | E0434 | PORTABLE LIQUID OXYGEN SYSTEM, RENTAL; INCLUDES PORTABLE CONTAINER, SUPPLY RESERVOIR, HUMIDIFIER, FLOWMETER, REFILL ADAPTOR, CONTENTS GAUGE, CANNULA OR MASK, AND TUBING |
| Supplemental<br>O2 | Procedure code | ICD-10-PCS | E0435 | PORTABLE LIQUID OXYGEN SYSTEM, PURCHASE; INCLUDES PORTABLE CONTAINER, SUPPLY RESERVOIR, FLOWMETER, HUMIDIFIER, CONTENTS GAUGE, CANNULA OR MASK, TUBING AND REFILL ADAPTOR |
|  |  | ICD-10-PCS | E0439 | STATIONARY LIQUID OXYGEN SYSTEM, RENTAL; INCLUDES CONTAINER, CONTENTS, REGULATOR, FLOWMETER, HUMIDIFIER, NEBULIZER, CANNULA OR MASK, & TUBING |
|  |  | ICD-10-PCS | E0440 | STATIONARY LIQUID OXYGEN SYSTEM, PURCHASE; INCLUDES USE OF RESERVOIR, CONTENTS INDICATOR, REGULATOR, FLOWMETER, HUMIDIFIER, NEBULIZER, CANNULA OR MASK, AND TUBING |
|  |  | ICD-10-PCS | E0441 | STATIONARY OXYGEN CONTENTS, GASEOUS, 1 MONTH'S SUPPLY = 1 UNIT |
|  |  | ICD-10-PCS | E0442 | STATIONARY OXYGEN CONTENTS, LIQUID, 1 MONTH'S SUPPLY = 1 UNIT |
|  |  | ICD-10-PCS | E0443 | PORTABLE OXYGEN CONTENTS, GASEOUS, 1 MONTH'S SUPPLY = 1 UNIT |
|  |  | ICD-10-PCS | E0444 | PORTABLE OXYGEN CONTENTS, LIQUID, 1 MONTH'S SUPPLY = 1 UNIT |
|  |  | ICD-10-PCS | E0455 | OXYGEN TENT, EXCLUDING CROUP OR PEDIATRIC TENTS |
|  |  | ICD-10-PCS | E1353 | REGULATOR |
|  |  | ICD-10-PCS | E1355 | STAND/RACK |
|  |  | ICD-10-PCS | E1390 | OXYGEN CONCENTRATOR, SINGLE DELIVERY PORT, CAPABLE OF DELIVERING 85 PERCENT OR GREATER OXYGEN CONCENTRATION AT THE PRESCRIBED FLOW RATE |
|  |  | ICD-10-PCS | E1391 | OXYGEN CONCENTRATOR, DUAL DELIVERY PORT, CAPABLE OF DELIVERING 85 PERCENT OR GREATER OXYGEN CONCENTRATION AT THE PRESCRIBED FLOW RATE, EACH |
|  |  | ICD-10-PCS | E1392 | PORTABLE OXYGEN CONCENTRATOR, RENTAL |
|  |  | ICD-10-PCS | E1405 | OXYGEN AND WATER VAPOR ENRICHING SYSTEM WITH HEATED DELIVERY |
|  |  | ICD-10-PCS | E1406 | OXYGEN AND WATER VAPOR ENRICHING SYSTEM WITHOUT HEATED DELIVERY |
|  |  | ICD-10-PCS | K0671 | PORTABLE OXYGEN CONCENTRATOR, RENTAL |
|  |  | ICD-10-PCS | K0738 | PORTABLE GASEOUS OXYGEN SYSTEM, RENTAL; HOME COMPRESSOR USED TO FILL PORTABLE OXYGEN CYLINDERS; INCLUDES PORTABLE CONTAINERS, REGULATOR, FLOWMETER, HUMIDIFIER, CANNULA OR MASK, AND TUBING |
|  |  | ICD-10-PCS | K0740 | REPAIR OR NONROUTINE SERVICE FOR OXYGEN EQUIPMENT REQUIRING THE SKILL OF A TECHNICIAN, LABOR COMPONENT, PER 15 MINUTES |
|  |  | ICD-10-PCS | K0741 | PORTABLE GASEOUS OXYGEN SYSTEM, RENTAL, INCLUDES PORTABLE CONTAINER, REGULATOR, FLOWMETER, HUMIDIFIER, CANNULA OR MASK, AND TUBING, FOR CLUSTER HEADACHES |
|  |  | ICD-10-PCS | QH | OXYGEN CONSERVING DEVICE IS BEING USED WITH AN OXYGEN DELIVERY SYSTEM |
|  |  | ICD-10-PCS | S8120 | OXYGEN CONTENTS, GASEOUS, 1 UNIT EQUALS 1 CUBIC FOOT |
|  |  | ICD-10-PCS | S8121 | OXYGEN CONTENTS, LIQUID, 1 UNIT EQUALS 1 POUND |
|  | Revenue Code | RevCode | 277 | Medical/surgical supplies-oxygen-take home |
|  |  | RevCode | 413 | Respiratory services-hyperbaric oxygen therapy |
|  |  | RevCode | 544 | Ambulance-oxygen |
|  |  | RevCode | 600 | Oxygen-general classification |
|  |  | RevCode | 601 | Oxygen-stat or port equip/supply or count |
|  |  | RevCode | 602 | Oxygen-stat/equip/under 1 LPM |
|  |  | RevCode | 603 | Oxygen-stat/equip/over 4 LPM |
|  |  | RevCode | 604 | Oxygen-stat/equip/portable add-on |
| Supplemental O2 | Dx indicating procedure | ICD-10-CM | Z99.81 | Dependence on supplemental oxygen |
|  | Dx indicating clinical need for procedure | ICD-10-CM | J96.91 | Respiratory failure, unspecified with hypoxia |
|  |  | ICD-10-CM | J96.01 | Acute respiratory failure with hypoxia |
|  |  | ICD-10-CM | J96.11 | Chronic respiratory failure with hypoxia |
|  |  | ICD-10-CM | J96.21 | Acute and chronic respiratory failure with hypoxia |
|  |  | ICD-10-CM | R09.0 | Asphyxia and hypoxemia |
|  |  | ICD-10-CM | R09.02 | Hypoxemia |

**Table A.3.**
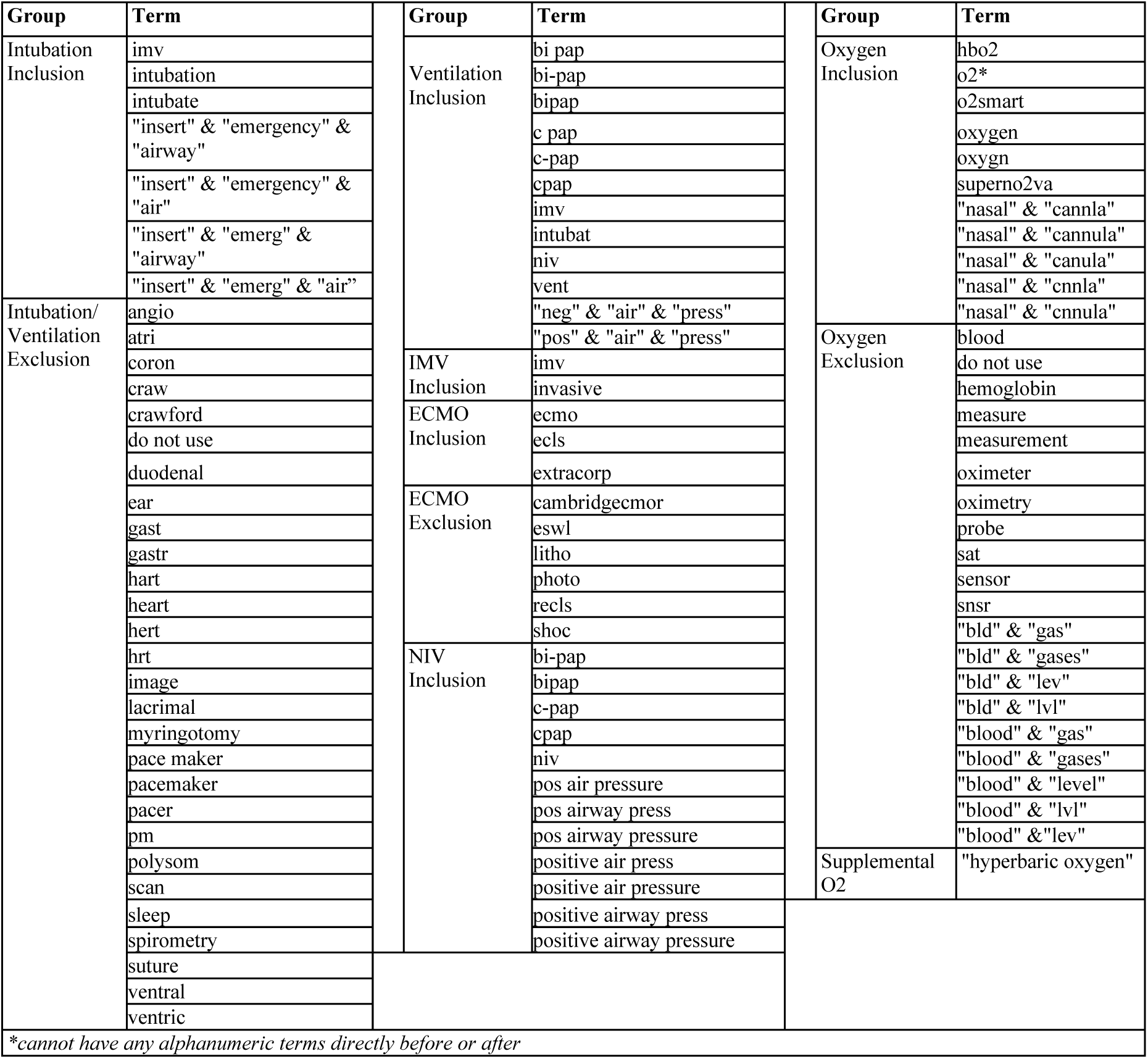
Chargemaster charge code and departmental code lists for the expanded-administrative algorithm for use in HealthVerity data (search terms may require modification for other administrative real-world data sources)

## APPENDIX B: Additional Results

**Table B.1.** Attrition of Optum and HealthVerity data.

|  | Optum de-identified<br>COVID-19 EHR<br>(N=2,018,728) |  | HealthVerity<br>administrative data<br>(N=35,629,662) |  |
| --- | --- | --- | --- | --- |
|  | % Excluded | N Remaining | % Excluded | N Remaining |
| Patients hospitalized for COVID-19 |  | 30,134 |  | 53,077 |
| Activity in baseline (183 days) | (-31.6%) | 20,599 | (-11.4%) | 47,048 |
| No missing age, sex, or region | (-13.3%) | 17,853 | (-2.4%) | 45,937 |
| No prior CSI during new user washout (90d) | (-13.0%) | 15,527 | (-18.6%) | 37,379 |
| **DEX+ vs. CSI-, initial algorithm** |  | **1,768 pairs**<br>**(3,536 total)** |  | **5,524 pairs**<br>**(11,048 total)** |
| **NEITHER** pairs |  | 1,437 (81.3%) |  | 3,615 (65.4%) |
| **O2/NIV** pairs |  | 293 (16.6%) |  | 1,644 (29.8%) |
| **IMV** pairs |  | 38 (2.1%) |  | 265 (4.8%) |
| **DEX+ vs. CSI-, expanded algorithm** |  | **1,757 pairs**<br>**(3,514 total)** |  | **5,337 pairs**<br>**(10,674 total)** |
| **NEITHER** pairs |  | 591 (33.6%) |  | 1,705 (31.9%) |
| **O2/NIV** pairs |  | 1,045<br>(59.5%) |  | 3,234 (60.6%) |
| **IMV** pairs |  | 121 (6.9%) |  | 398 (7.5%) |

**Table B.2.**
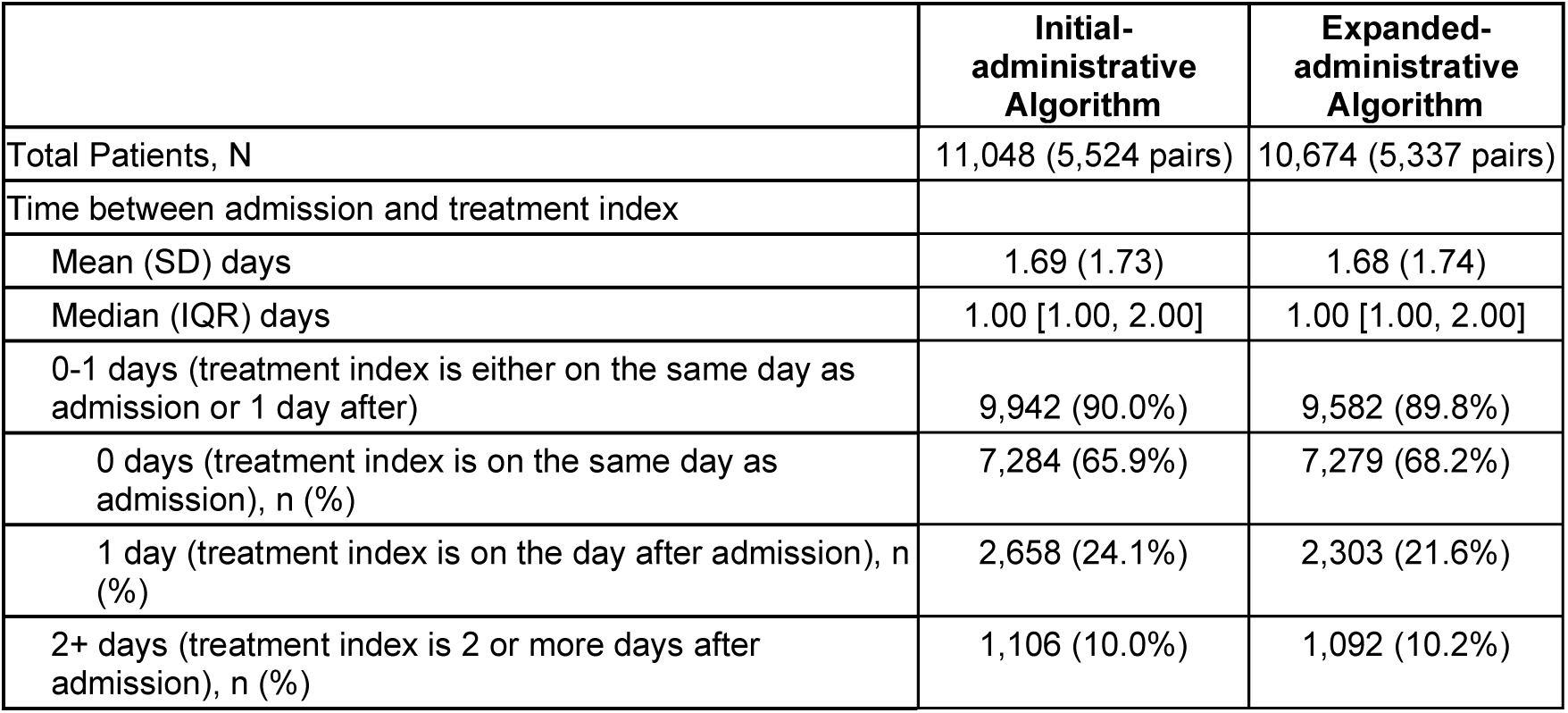
Time between admission and treatment index among patients in the administrative data cohort matched using the initial-administrative and the expanded-administrative algorithms.

**Figure B.1.**
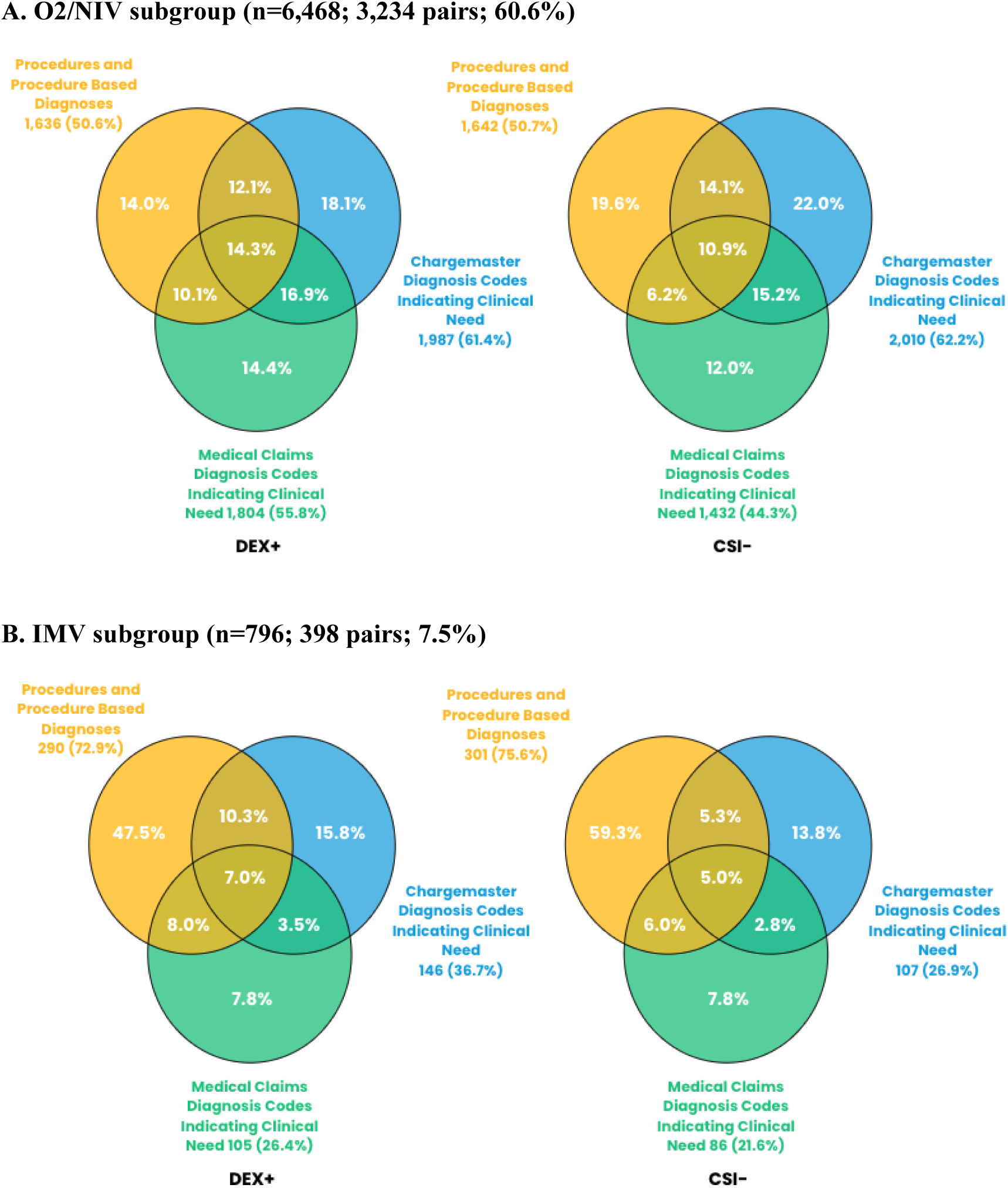
Measure component contributions to the mWHO severity score in each treatment arm of the administrative data cohort matched using the expanded-administrative algorithm (n=10,674; 5,337 pairs)

**Figure B.2.**
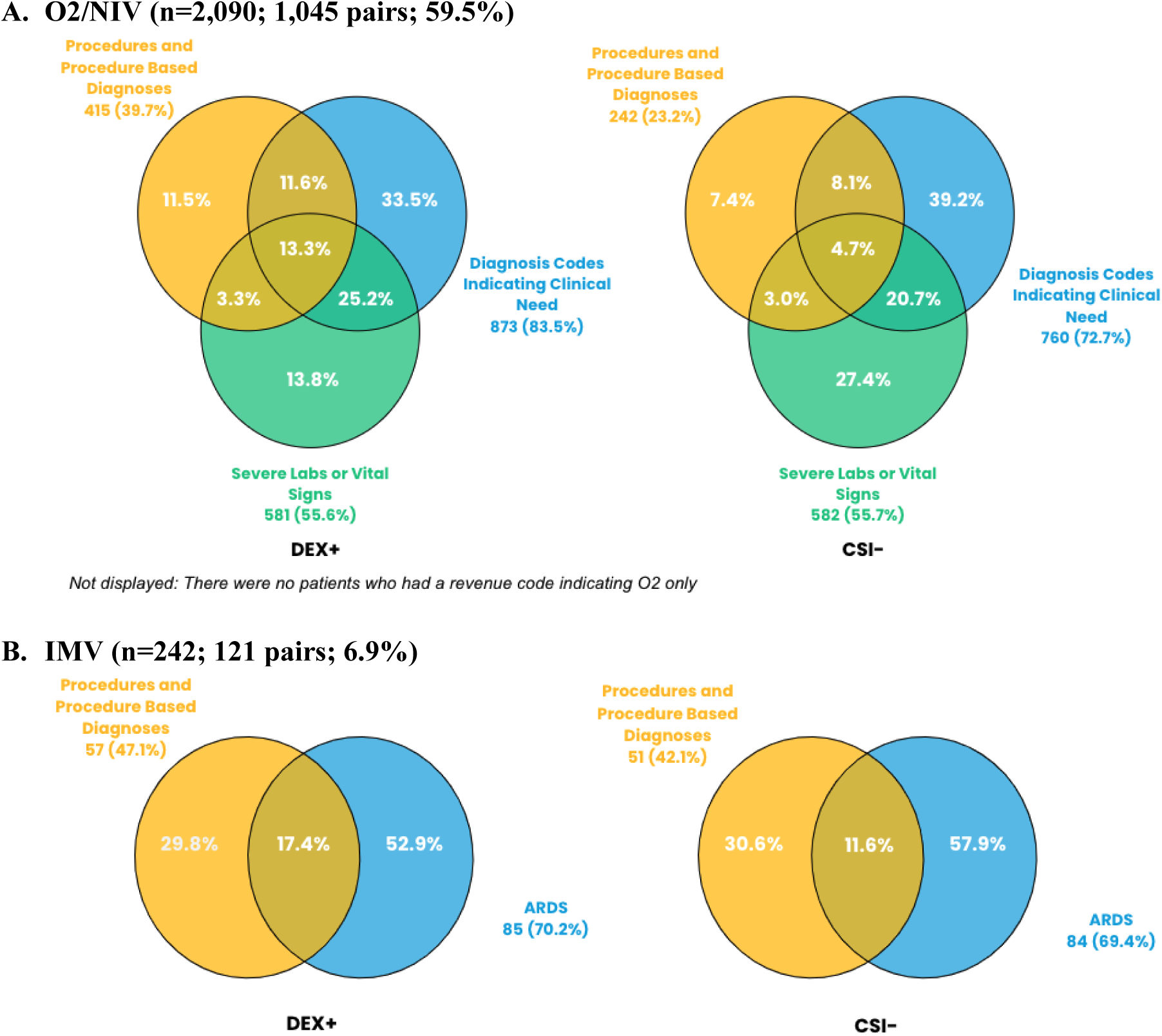
Drivers of the shift from lower to greater COVID-19 severity in each treatment arm in the EHR-based cohort matched using the expanded-EHR algorithm (n=3,514; 1,757 pairs)

**Figure B.3.**
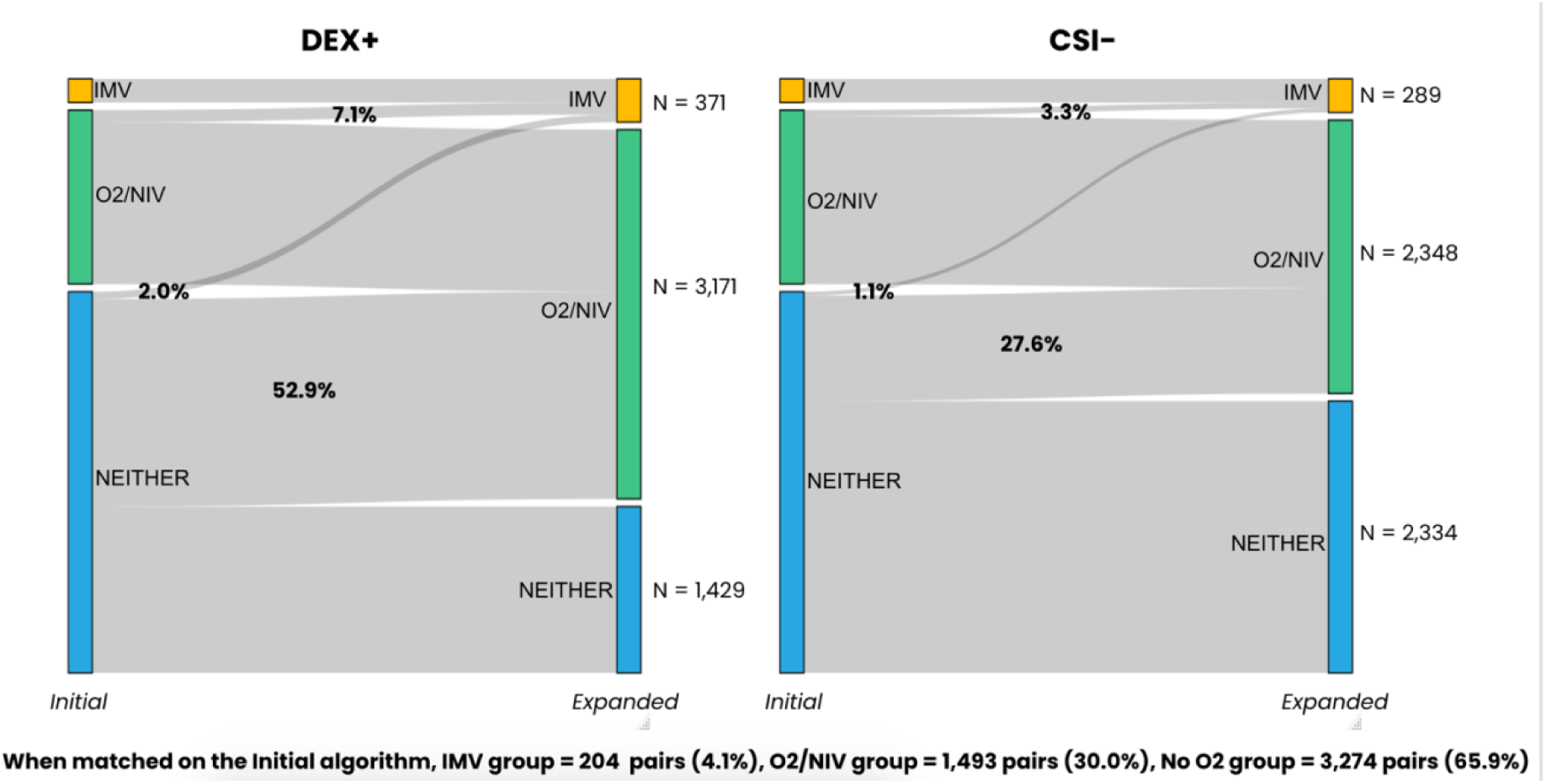
Sensitivity analysis: Shifts in COVID-19 severity categories in each treatment arm in the administrative data cohort matched using the initial-administrative algorithm among the subset of patients who initiated treatment on the same day or 1 day after the admission date (n=9,942; 4,971 pairs)

**Figure B.4.**
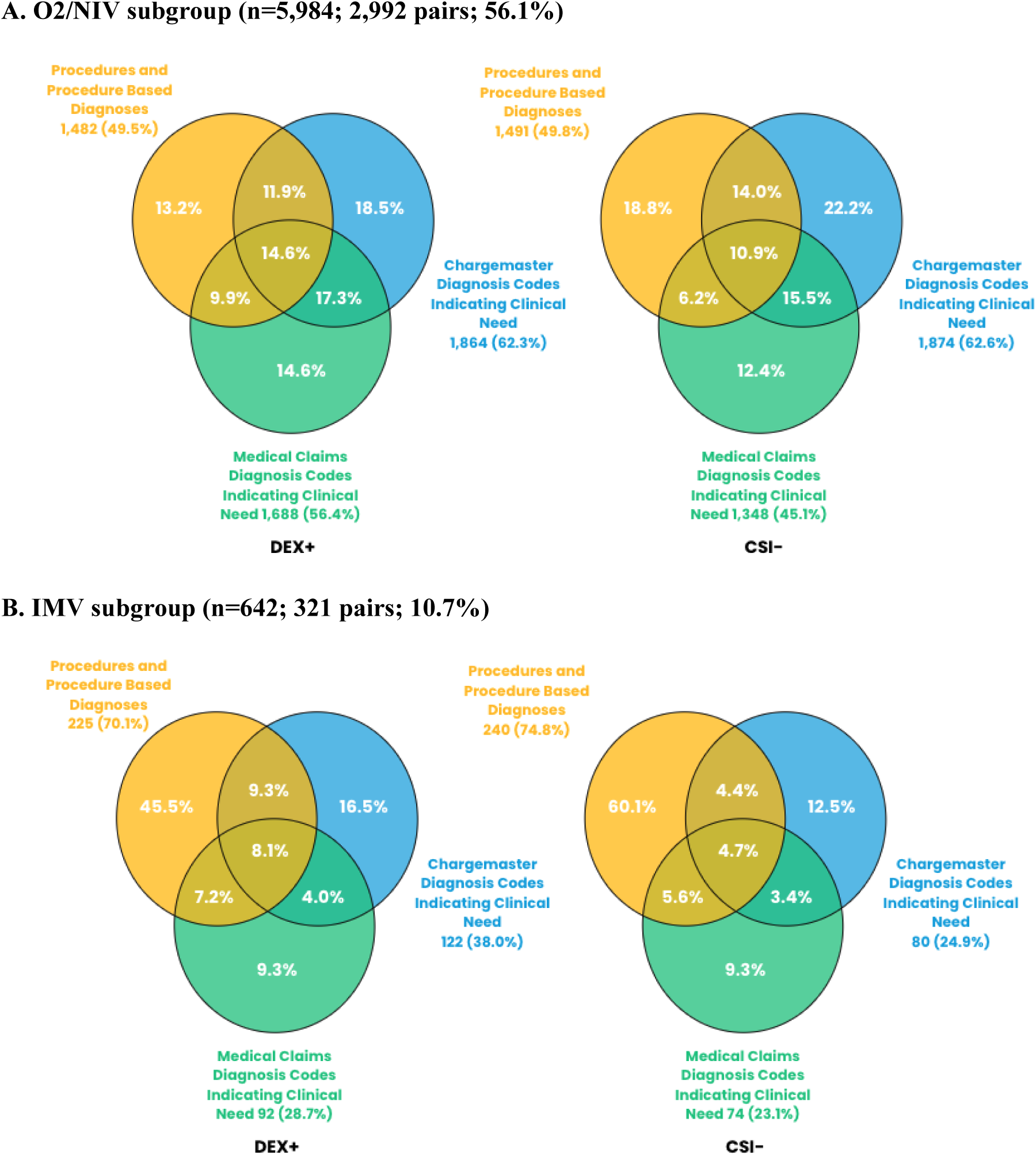
Sensitivity analysis: Measure component contributions to the mWHO severity categorization in each treatment arm of the administrative data cohort matched using the expanded-administrative algorithm among the subset of patients who initiated treatment on the same day or 1 day after the admission date (n=10,674; 5,337 pairs)

## APPENDIX C. Additional Data Source Detail

### C.1 HealthVerity Data

The HealthVerity COVID-19 data includes 16 unique US data sources from all 50 states across four main data types (described in Table C.1 and illustrated in Figure C.1). Patients with at least one medical and pharmacy claim, and any available CDM, EHR, and lab data are linked across data types using a unique patient identifier. Data capture began December 2018, with the exception of two lab sources that began in March/June 2020. The data is refreshed approximately every 2 weeks with varying data lag by data type and vendor (see Table C.1). Additional specifications of the HealthVerity COVID-19 dataset are summarized in Table C.2.

**Table C.1.** HealthVerity description of each data type and data lag/refresh rate.

| Data type | Description | Data lag/refresh rate |
| --- | --- | --- |
| **Medical and pharmacy claims**<br>(both open and closed claims) | Medical insurance claims and outpatient pharmacy transactions are sourced from provider-submitted claims, adjudicated insurance claims, and pharmacy benefit manager billing (nine sources in total). Open claims are sourced from practice management systems, billing systems and claims clearinghouses, while closed claims are sourced from insurance providers and payers. Data encompass all major payer types (commercial, Medicaid, Medicare), and include details on service dates, medications, diagnoses, and procedures. | Open claims data are sourced in near-real-time with a lag by two weeks, while closed claims may lag by a month or more but offer a comprehensive and longitudinal history in a closed ecosystem (follow-up is 12+ months for most patients). Across the nine claims sources, the data lags range from 1-90 days, and are refreshed weekly. |
| **Chargemaster**<br>(CDM, formerly known as charge description master) | Sourced from administrative hospital billing systems, chargemaster data capture all drugs, procedures, and medical services provided during a hospital stay. Drug, procedure, and medical data are day-level captured, and admitting and primary diagnoses are available for each patient encounter. Outpatient hospital data (e.g., outpatient visits taking place at an ambulatory medical center) are also captured. | Data lag per patient discharge, with data released typically within 10 days of discharge. The data are refreshed monthly. |
| **Electronic health records**<br>(EHR) | Select clinical observations from EHR sources (e.g., BMI, smoking status) augment the claims and CDM data, and allow for further analysis by population subgroup. | The data lags 1-2.5 months, and are refreshed monthly. |
| **Labs** | Lab testing activity and results data, including for COVID-19 NAAT/PCR and serology tests, are available. Sources include Quest Diagnostics and other nationally-representative laboratories. | The data lags by two weeks, and are refreshed weekly. |

**Figure C.1.**
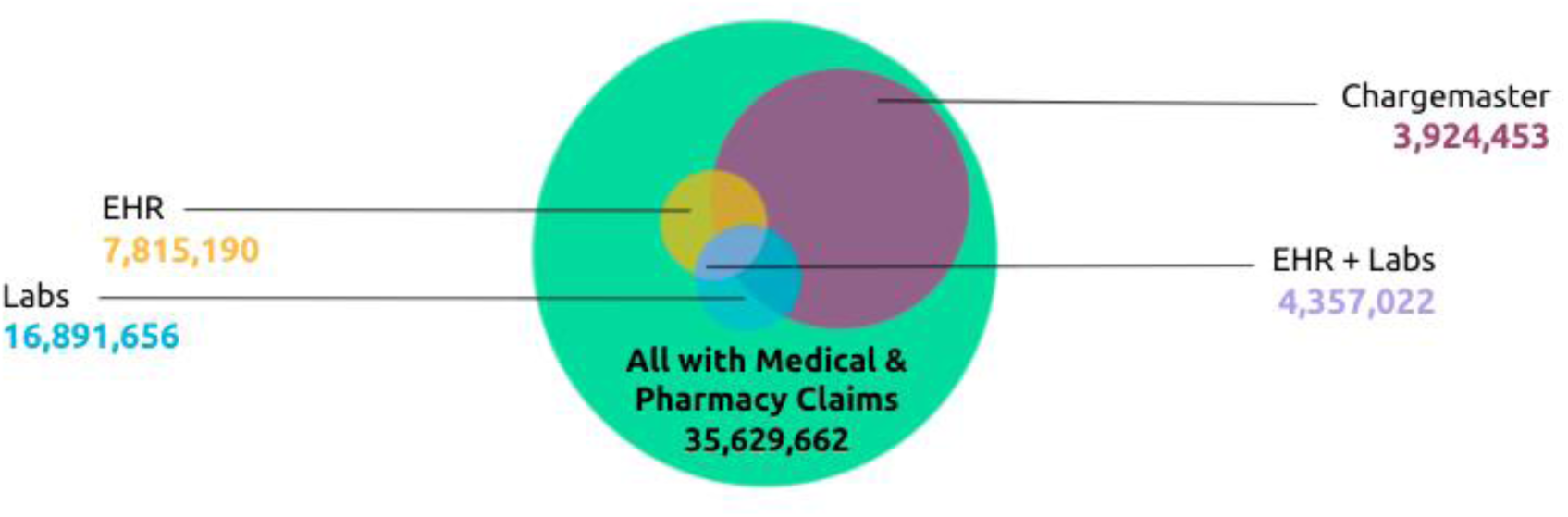
Illustration of data type overlap among HealthVerity patients with medical and pharmacy claims as of August 31, 2020 (N=35,629,662)

**Table C.2.**
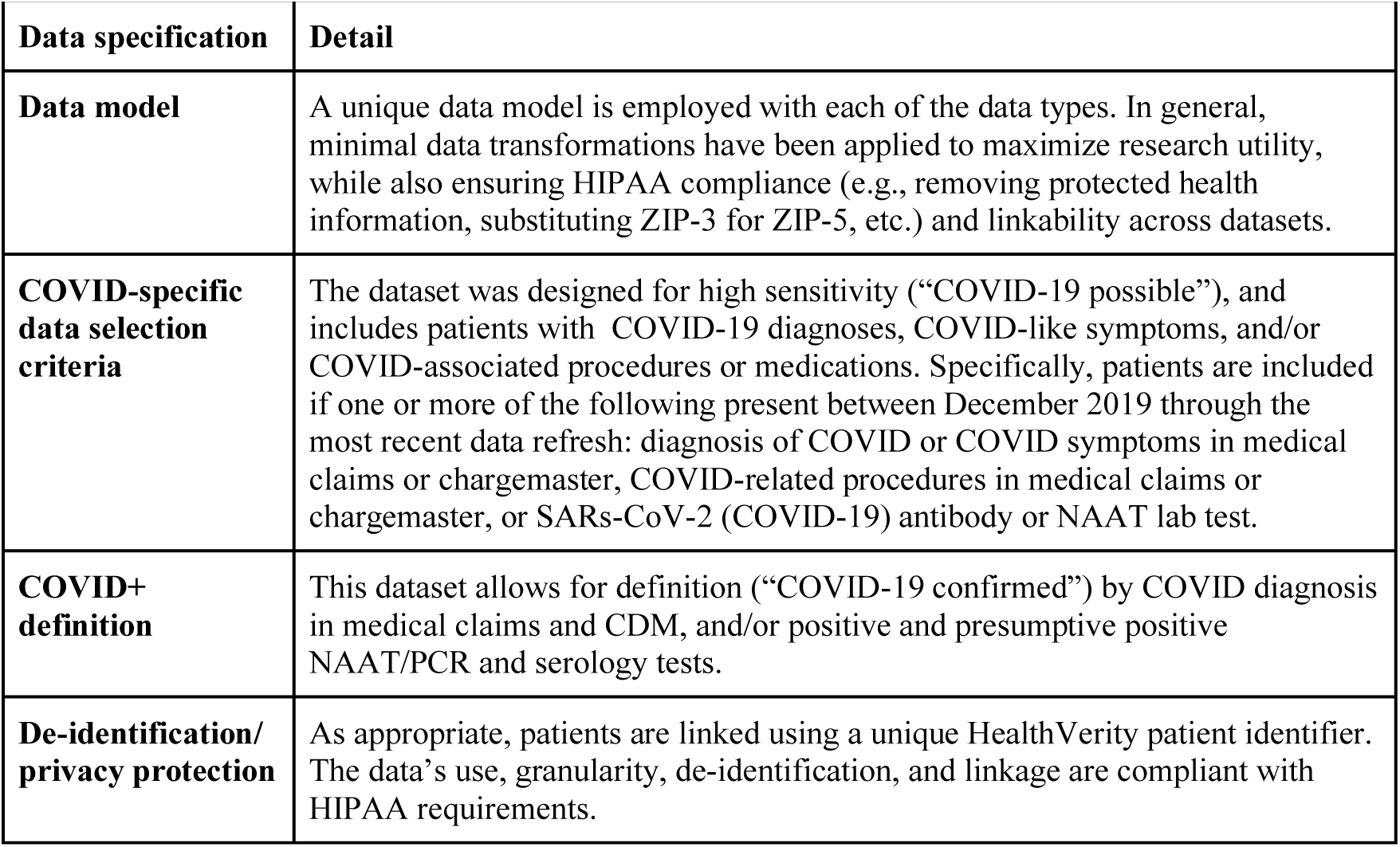
HealthVerity overview of data specifications.

| Data specification | Detail |
| --- | --- |
| **Data model** | A unique data model is employed with each of the data types. In general, minimal data transformations have been applied to maximize research utility, while also ensuring HIPAA compliance (e.g., removing protected health information, substituting ZIP-3 for ZIP-5, etc.) and linkability across datasets. |
| **COVID-specific data selection criteria** | The dataset was designed for high sensitivity (“COVID-19 possible”), and includes patients with COVID-19 diagnoses, COVID-like symptoms, and/or COVID-associated procedures or medications. Specifically, patients are included if one or more of the following present between December 2019 through the most recent data refresh: diagnosis of COVID or COVID symptoms in medical claims or chargemaster, COVID-related procedures in medical claims or chargemaster, or SARs-CoV-2 (COVID-19) antibody or NAAT lab test. |
| **COVID+ definition** | This dataset allows for definition (“COVID-19 confirmed”) by COVID diagnosis in medical claims and CDM, and/or positive and presumptive positive NAAT/PCR and serology tests. |
| **De-identification/privacy protection** | As appropriate, patients are linked using a unique HealthVerity patient identifier. The data’s use, granularity, de-identification, and linkage are compliant with HIPAA requirements. |

### C.2 Optum COVID-19 De-identified Electronic Health Records

The Optum COVID-19 de-identified electronic health records (EHR) is sourced from laboratories and hospital and emergency department EHRs from integrated delivery networks (IDNs) and smaller outpatient clinics from all over the country. The data in the analysis is entirely inpatient and includes diagnosis data, laboratory data with results, procedures, vital sign measurements, prescriptions written, and medications administered. Sourced from the legacy Humedica database, now Optum EHR, the limited dataset includes a subset of patients as described in the COVID-specific data selection criteria of Table C.3. Data capture began February 1, 2020 and ended on September 24, 2020 with no scheduled updates and includes approximately 2 million patients (N=2,018,728). If patients were already in the Optum EHR database, patient history was included. The underlying data is representative of the US, but the COVID-19 cut of data is skewed towards the Midwest and Northeast. Additional specifications of the Optum COVID-19 dataset are summarized in Table C.3.

**Figure C.2.**
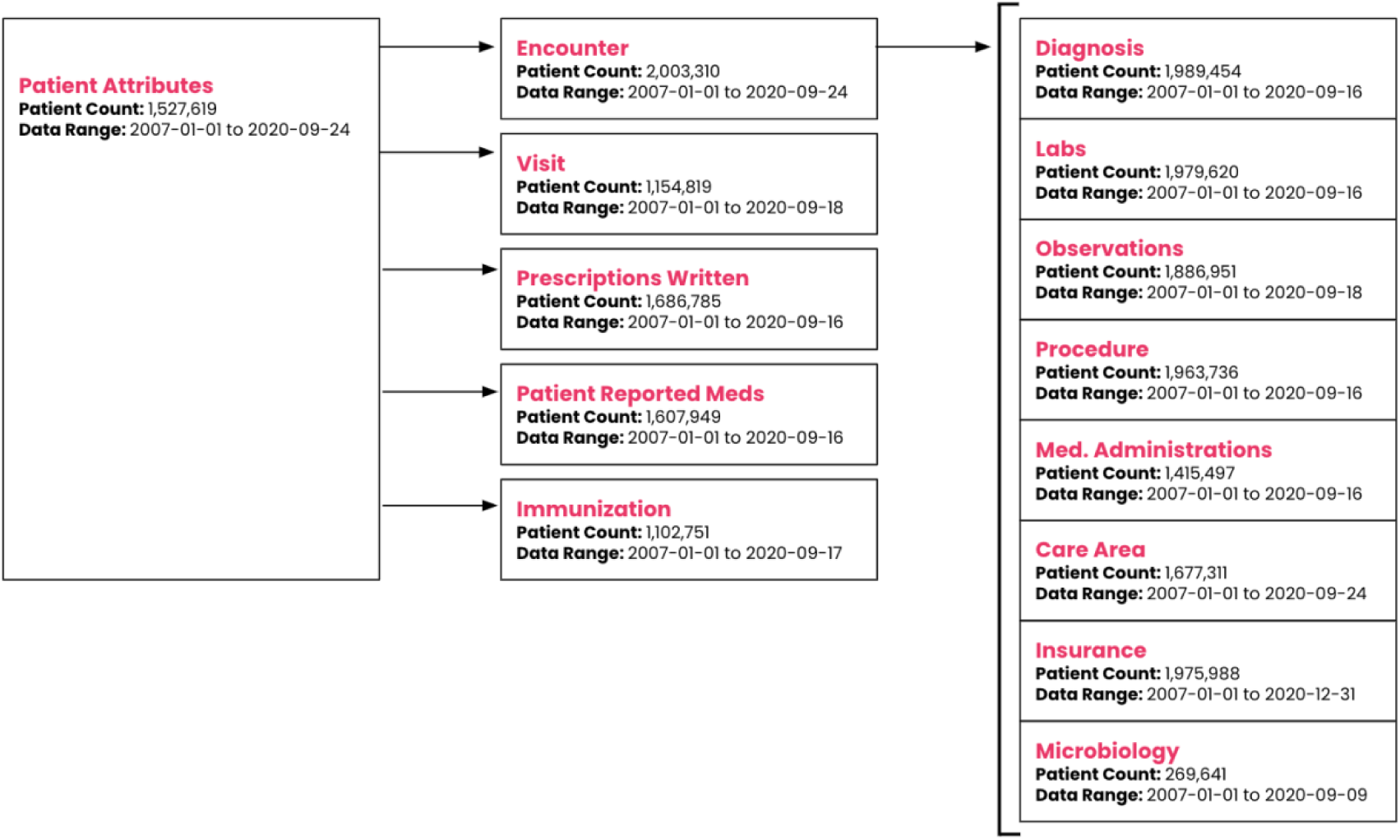
Optum COVID-19 Data Schema and Summary of Patient Counts and Event-Specific Data Ranges.

**Table C.3.** Optum COVID-19 EHR overview of data specifications.

| **Data specification** | **Detail** |
| --- | --- |
| **Data model** | Datasets are merged from several EHRs. To complete the timeliness of this data, Optum may impute information to help with this merge. The data are HIPAA-compliant while preserving data integrity. Optum EHR has a unique to themselves data model. |
| **COVID-specific data selection criteria** | Data for a select set of patients are pulled from the broader Optum EHR dataset and included in the COVID dataset if they are COVID “possible”, according to the following data elements.<br>**ICD-10 diagnosis codes:**<br>COVID-19 diagnosis (*B97.29, B34.2, U07.1, U07.2*); influenza-like illness, including acute bronchitis, lower respiratory infection (*B97.29, J12.89, J20.8, J22\*, J40\*, J98.8*); exposure to COVID-19 (*Z20.828*); cough (*R05.\**); shortness of breath (*R06.02*); fever (*R50.9*); acute respiratory distress syndrome (*J80.\**); screening for COVID-19 (*Z11.59*)<br>**CPT/HCPCS:**<br>SARs-CoV-2 (COVID-19) lab test procedures (*86328, 86769, 87635, G2023, G2024, U0001, U0002, U0003, U0004*)<br>**LOINC (Logical Observation Identifiers Names and Codes):**<br>SARs-CoV-2 (COVID-19) lab test orders (*94534-5, 94500-6, 94503-0, 94505-5, 94506-3, 94504-8, 94507-1, 94509-7, 94508-9, 94510-5, 94308-4, 94309-2, 94311-8, 94312-6, 94314-2, 94315-9, 94316-7, 94533-7, 94531-1, 94547-7, 94564-2, 94562-6, 94563-4, 94565-9, 94307-6, 94559-2, 94306-8, 94558-4, 94511-3*)<br>**Lab Test Name:**<br>Text search corresponding to SARs-CoV-2 (COVID-19) lab test procedures/orders included in the CPT/HCPCS and LOINC lists above |
| **COVID+ definition** | This dataset allows for definition (“COVID-19 confirmed”) by COVID diagnosis in the EHR, and/or positive and presumptive positive NAAT/PCR and serology tests from the labs table. |
| **De-identification/ privacy protection** | Optum uses Datavant as their de-identification engine. All data is HIPAA compliant. |

## References

1. Barrot L, Asfar P, Mauny F, et al. Liberal or Conservative Oxygen Therapy for Acute Respiratory Distress Syndrome. New England Journal of Medicine. 2020;382(11):999–1008. doi:10.1056/NEJMoa1916431

2. Burn E, Sena AG, Prats-Uribe A, et al. Use of dialysis, tracheostomy, and extracorporeal membrane oxygenation among 240,392 patients hospitalized with COVID-19 in the United States. medRxiv [Preprint]. 2020 Nov 27:2020.11.25.20229088. Doi: 10.1101/2020.11.25.20229088. PMID: 33269356; PMCID: PMC7709172.

3. Centers for Disease Control (CDC). COVIDView: A weekly surveillance summary of U.S. Covid-19 activity. Cdc.gov. Published December 11, 2020. Accessed May 14, 2021. https://www.cdc.gov/coronavirus/2019-ncov/covid-data/covidview/index.html

4. Chu DK, Kim LH-Y, Young PJ, et al. Mortality and morbidity in acutely ill adults treated with liberal versus conservative oxygen therapy (IOTA): a systematic review and meta-analysis. Lancet. 2018;391(10131):1693–1705. doi:10.1016/S0140-6736(18)30479-3.

5. Fan E, Brodie D, Slutsky AS. Acute Respiratory Distress Syndrome: Advances in Diagnosis and Treatment. JAMA. 2018;319(7):698–710. doi:10.1001/jama.2017.21907

6. Gordon DE, Hiatt J, Bouhaddou M, et al. Comparative host-coronavirus protein interaction networks reveal pan-viral disease mechanisms. Science. 2020 Dec 4;370(6521):eabe9403. doi:10.1126/science.abe9403. Epub 2020 Oct 15. PMID: 33060197.

7. Food and Drug Administration (FDA). COVID-19: Developing Drugs and Biological Products for Treatment or Prevention Guidance for Industry. Updated February 22, 2021. https://www.fda.gov/regulatory-information/search-fda-guidance-documents/covid-19-developing-drugs-and-biological-products-treatment-or-prevention

8. Harvey RA, Rassen JA, Kabelac CA, Turenne W, Leonard S, Klesh R, Meyer WA 3rd, Kaufman HW, Anderson S, Cohen O, Petkov VI, Cronin KA, Van Dyke AL, Lowy DR, Sharpless NE, Penberthy LT. Association of SARS-CoV-2 Seropositive Antibody Test With Risk of Future Infection. JAMA Intern Med. 2021 Feb 24. doi: 10.1001/jamainternmed.2021.0366. Epub ahead of print. PMID: 33625463.

9. Hughes R, Whitley L, Fitovski K, Schneble HM, Muros E, Sauter A, Craveiro L, Dillon P, Bonati U, Jessop N, Pedotti R, Koendgen H. COVID-19 in ocrelizumab-treated people with multiple sclerosis. Mult Scler Relat Disord. 2020 Dec 30;49:102725. doi: 10.1016/j.msard.2020.102725. Epub ahead of print. PMID: 33482590; PMCID: PMC7772086. https://www.ncbi.nlm.nih.gov/pmc/articles/PMC7772086/

10. Murk W, Gierada M, Fralick M, et al. Diagnosis-wide analysis of COVID-19 complications: an exposure-crossover study. CMAJ. 2020 Dec 7;193(1):E10–8. doi: 10.1503/cmaj.201686. Epub ahead of print. PMID: 33293424; PMCID: PMC7774475.

11. National Institute of Health (NIH). Coronavirus Disease 2019 (COVID-19) Treatment Guidelines. Updated March 5, 2021. https://www.covid19treatmentguidelines.nih.gov/whats-new/

12. Ni Y-N, Luo J, Yu H, Liu D, Liang B-M, Liang Z-A. The effect of high-flow nasal cannula in reducing the mortality and the rate of endotracheal intubation when used before mechanical ventilation compared with conventional oxygen therapy and noninvasive positive pressure ventilation. A systematic review and meta-analysis. Am J Emerg Med. 2018;36(2):226–233. doi:10.1016/j.ajem.2017.07.083

13. Papazian L, Aubron C, Brochard L, et al. Formal guidelines: management of acute respiratory distress syndrome. Ann Intensive Care. 2019;9. doi:10.1186/s13613-019-0540-9

14. Quan H, Sundararajan V, Halfon P, Fong A, Burnand B, Luthi JC, Saunders LD, Beck CA, Feasby TE, Ghali WA. Coding algorithms for defining comorbidities in ICD-9-CM and ICD-10 administrative data. Med Care. 2005 Nov;43(11):1130–9. doi:10.1097/01.mlr.0000182534.19832.83. PMID: 16224307.

15. RECOVERY Collaborative Group, Horby P, Lim WS et al. Dexamethasone in Hospitalized Patients with Covid-19 - Preliminary Report. N Engl J Med. 2020 Jul 17:NEJMoa2021436. doi: 10.1056/NEJMoa2021436. Epub ahead of print. PMID: 32678530; PMCID: PMC7383595. https://www.ncbi.nlm.nih.gov/pmc/articles/PMC7383595/

16. Rochwerg B, Brochard L, Elliott MW, et al. Official ERS/ATS clinical practice guidelines: noninvasive ventilation for acute respiratory failure. European Respiratory Journal. 2017;50(2). doi:10.1183/13993003.02426-2016

17. Sievert C. Interactive Web-Based Data Visualization with R, plotly, and shiny. Chapman and Hall/CRC Florida, 2020.

18. Wang SV, et al. Transparency and Reproducibility of Observational Cohort Studies Using Large Healthcare Databases. Clin Pharmacol Ther. 2016 Mar;99(3):325–332. doi: 10.1002/cpt.329

19. Wickham H, Averick M, Bryan J, et al. Welcome to the Tidyverse. J Open Source Softw. 2019;4(43):1686. doi:10.21105/joss.01686.

20. World Health Organization (WHO). Coronavirus Disease (COVID-19) Dashboard. Geneva: World Health Organization, 2020. Available online: https://covid19.who.int/. Accessed May 14, 2021.

21. World Health Organization (WHO) Working Group on the Clinical Characterisation and Management of COVID-19 infection. A minimal common outcome measure set for COVID-19 clinical research. Lancet Infect Dis. 2020 Aug;20(8):e192–e197. doi: 10.1016/S1473-3099(20)30483-7. Epub 2020 Jun 12. Erratum in: Lancet Infect Dis. 2020 Oct;20(10):e250. PMID: 32539990; PMCID: PMC7292605. https://www.ncbi.nlm.nih.gov/pmc/articles/PMC7292605/

22. Yih WK, Hua W, Draper C, Dutcher S, Fuller C, Kempner M, Kit B, Lyons J, Driscoll MR, Toh SD, Lo Re V. COVID-19 Natural History Master Protocol. Sentinel Initiative, version 3.0, October 9, 2020. https://www.sentinelinitiative.org/sites/default/files/Methods/COVID-19_Natural_History_Protocol_V3.0.pdf

